# Rare predicted loss-of-function and damaging missense variants in *CFHR5* associate with protection from age-related macular degeneration

**DOI:** 10.1101/2024.11.13.24317290

**Authors:** Aaron M. Holleman, Aimee M. Deaton, Rachel A. Hoffing, Lynne Krohn, Philip LoGerfo, Paul Nioi, Mollie E. Plekan, Sebastian Akle Serrano, Simina Ticau, Tony E. Walshe, Anna Borodovsky, Lucas D. Ward

## Abstract

Age-related macular degeneration (AMD) is a leading cause of blindness among older adults worldwide, but treatment options are limited. Prior genetics studies have implicated the *CFH* locus, which contains *CFH* and the five *CFHR1-5* genes, in AMD. While the *CFH* gene has been robustly linked with AMD risk, the potential additional role of the five *CFHR* genes remains unclear, with strong linkage disequilibrium across the locus hindering the identification of individual gene contributions. Investigation of rare coding variants can help to identify causal genes in such regions. We used whole exome sequencing data from 406,952 UK Biobank participants to examine AMD associations with genes at the *CFH* locus. For each gene, we used burden testing to examine the association of rare (MAF<1%) predicted loss-of-function (pLOF) and predicted-damaging missense variants with AMD. When adjusting for *CFH*-region variants previously established to independently associate with AMD, we find that *CFHR5* rare variant burden significantly associates with decreased risk of AMD (OR=0.75, p=7×10^−4^), and that this association is primarily driven by pLOF variants. Furthermore, the association of *CFHR5* rare variants with AMD protection is estimated as stronger for individuals carrying the *CFH* Y402H variant, which increases AMD risk (interaction p=0.04). We also identify increased thinning of the outer segment of the photoreceptor layer of the retina to be a strong predictor of AMD and find that *CFHR5* rare variant burden significantly associates with increased thickness of this retinal layer (+0.34 SD, p=4×10^−4^). These findings suggest *CFHR5* inhibition as a potential therapeutic approach for AMD.

## INTRODUCTION

Age-related macular degeneration (AMD) is a progressive retinal disorder that impacts the macula, the central area of the retina responsible for fine vision. AMD has an estimated global prevalence of 8.7% among individuals 45 to 85 years old, and is the leading cause of blindness in developed countries.^1^ During early/intermediate stages of AMD, lipid- and protein-rich deposits called drusen accumulate, in tandem with pigmentary changes to the retinal pigmented epithelium (RPE).^2^ Progression to late-stage AMD can lead to RPE and photoreceptor cell death (dry form), and/or aberrant neovascularization of choroidal vessels from the sub-RPE space into the retina (wet form).^2^ Considering all AMD stages, dry-form AMD accounts for 85-90% of all AMD.^1^ Late-stage, vision-threatening AMD manifests with similar prevalences for dry and wet AMD forms.^1,3,4^

Anti-VEGF therapeutics delivered by intravitreal injection are the current standard of care for wet AMD. These therapies are effective at reversing fluid accumulation within the retina and can improve vision of wet AMD patients.^5^ However, these patients remain at risk for vision loss due to dry AMD, with 98% of individuals found to have macular atrophy after seven years of anti-VEGF treatment.^6^ Dry AMD therapeutics targeting components of the complement pathway have recently gained approval in the U.S., but these treatments do not improve vision, rather they slow the rate of atrophy over time, and like anti-VEGF therapies, can be burdensome as they require routine intravitreal administration.^7^ Hence, there is a continued need for novel treatments for both AMD subtypes, and a particularly large unmet need for dry AMD.

Genetics is a strong risk factor for AMD, with twin-based heritability estimated to be as high as 71%.^8^ In one of the first successful genome-wide association studies (GWAS) of a complex disease, Klein et al. detected a very strong AMD signal at the *CFH* locus, which contains the *CFH* gene and five *CFHR1-5* genes, and they linked this signal to the *CFH* Y402H missense variant.^9^ Simultaneously, two independent research groups using targeted genomic approaches also identified *CFH* Y402H as strongly increasing AMD risk.^10,11^ Following this early discovery, additional signals proximal to the *CFH* gene were identified to associate with AMD independently of the Y402H variant, including a common deletion of both *CFHR1* and *CFHR3* (Δ*CFHR3*-*CFHR1*) that associates with protection from AMD.^12,13^ In one of the largest GWAS of AMD to date, Fritsche et al. identified eight conditionally independent AMD association signals across the *CFH* locus, further highlighting the importance of this genomic region in AMD and the allelic heterogeneity at the *CFH* locus (**Figure S1**).^14^

Though a role for the *CFH* gene in AMD has been established, potential additional roles for each of the five *CFHR* genes remain unclear. Identification of possible independent roles for the *CFHR*s has been complicated by high linkage disequilibrium (LD) that exists between AMD-associated variants at the *CFH* gene and other variants across the entire *CFH* locus. To disentangle AMD signals and more effectively examine potential roles for each *CFHR* gene, investigators have used gene-based testing of low-frequency (MAF<5%) and rare (MAF<1%) variants while adjusting for the eight independent *CFH*-region AMD GWAS signals.^14,15^ Findings have suggested possible roles for certain *CFHR*s, including *CFHR2* and *CFHR5*,^15^ yet have been unable to convincingly demonstrate such roles, largely due to a lack of sequencing-based genetic data and power limitations that have prevented a more complete and robust investigation of independent rare variant associations, including for protein truncating variants (PTVs) and other predicted-damaging variants.

Recently, studies have made use of optical coherence tomography (OCT) imaging to gain an understanding of retinal layer changes that associate with and precede AMD, and have identified a thinner photoreceptor layer to be the retinal phenotype most strongly predictive of incident AMD and also most significantly associated with AMD polygenic risk score.^16,17^ Rare variant analyses of such AMD-predictive quantitative retinal phenotypes can act as an important complement to AMD case-control analyses in the effort to understand possible contributions by the *CFHR* genes to AMD.

In this study, we used exome-sequencing data from up to 406,952 UK Biobank (UKB) participants to analyze associations of gene-based rare (MAF<1%) PTVs and other predicted-damaging variants with AMD and AMD-predictive retinal phenotypes, with an overarching goal of clarifying the role of the *CFHR* genes in AMD pathophysiology.

## METHODS

### The UK Biobank resource

The UK Biobank (UKB) study is a population-based cohort study that recruited approximately 500,000 participants from England, Wales and Scotland between 2006 and 2010.^18^ Participants were aged 40-69 years at the time of recruitment. Phenotype data includes quantitative biomarker and anthropometric measures; as well as disease diagnoses based on self-report, inpatient hospitalizations, a cancer registry, and death records (the latter three sources are accessed through the National Health Service [NHS]). For approximately half of UKB participants, disease diagnoses are also available through general practitioner (GP) records. In addition, about 16% of participants have quantitative retinal layer thickness measures generated from Optical Coherence Tomography (OCT) imaging,^19,20^ and about 10% of participants have proteomics measurements based on Olink assays.^21^ Genotype data generated from array-based imputation are available for nearly all UKB participants, and genotypes based on whole exome sequencing (WES) are available for nearly 470,000 participants.

The UKB study was approved by the NHS National Research Ethics Service and all participants provided written informed consent allowing use of study data for health-related research. Information about ethics oversight in the UKB study can be found at https://www.ukbiobank.ac.uk/ethics/. We accessed data from the UKB resource using applications 26041 and 65851.

### WES data generation and processing

DNA was extracted from whole blood and WES was carried out by Regeneron Genetics Center.^22,23^ Exome capture was performed using the IDT xGen Exome Research Panel v1.0, and sequencing was performed with the Illumina NovaSeq 6000 platform. Sequence reads were then processed through the ‘OQFE protocol’, which is a Functional Equivalence (FE) protocol that retains original quality scores in the CRAM files.^24^ Briefly, raw sequence reads were mapped to the GRCh38 reference in a deterministic manner using BWA-MEM, duplicate reads were marked with Picard 2.21.2, and the final OQFE CRAM files were compressed, without recalibration or binning of quality scores. Variants were called on each CRAM file using DeepVariant 0.10.0, yielding a genomic VCF (gVCF) for each sample. gVCFs for all samples were then aggregated and joint genotyping was performed using GLnexus, resulting in multi-sample VCFs (pVCFs) that included genotype calls for all samples.

We used bcftools^25^ to normalize indels and split multiallelic sites, and then applied genotype filters that set individual genotypes to missing if genotype quality (GQ) <20, or if read depth (DP) <7 for single nucleotide variants (SNVs) or DP<10 for indels. Picard^26^ was used to calculate alternate allele balance (AB) and we excluded variant sites that did not have ≥1 sample with AB≥0.15 for SNVs or AB≥0.20 for indels.

We then grouped participants according to the genetic ancestry assignments made by the Pan-UK Biobank study at the Broad Institute.^27^ We decided to restrict analyses to the European ancestry grouping, given the lack of power for identifying rare variant associations using the much smaller sample sizes of the other five ancestral groupings.

For UKB participants in the European ancestry grouping, we used PLINK2^28,29^ to apply sample quality control. Individuals whose self-reported gender and genetic-inferred sex were discordant and those with sex chromosome aneuploidy were removed. Using only the set of variants with WES genotype missingness <1%, we identified and excluded samples missing >1% of genotypes. We also used a set of high-quality, low-LD common variants to identify and remove samples with outlier heterozygosity estimates (F<-0.15 or F>0.15). For rare variant analyses, we retained only variants with genotypes missing for <5% of participants, those with Hardy-Weinberg equilibrium (HWE) mid-p-value >1×10^−12^, and those with MAF<1%.

### Generation of principal components for use as covariates

Following the analyses of Backman et al. (2021),^23^ we used a combination of principal components (PCs) generated from genome-wide common-variant data and PCs generated from exome-wide rare-variant data.

To generate PCs based on common variants, we used the genome-wide imputation-based genotypes that are available for nearly all UKB participants.^18^ For the set of unrelated participants with European genetic ancestry, we limited to autosomal SNPs (i.e., no indels) with INFO=1, missingness <2%, HWE mid-p-value >1×10^−12^, and MAF>1%. We removed genomic regions known to have long-range, high LD.^30^ Then we used PLINK2 to perform LD pruning using the flag *--indep-pairwise 1000 100 0*.*15*. Using this set of high-quality, independent, genome-wide SNPs, we ran smartpca^31^ to generate common-variant-based PCs, first estimating PCs using only unrelated participants, then projecting related participants onto the PCs.

Using the WES data, we also generated a set of PCs based on exome-wide rare variants, as prior studies have demonstrated that bias due fine-scale population structure can persist when controlling for PCs based on common variants only.^23^ This PC process mirrored the common-variant-based PC process described in the preceding paragraph, with the exception that the variants used for PC generation had MAF<1% and minor allele count (MAC) ≥100.

### Testing gene-based rare variant associations with ‘broadly defined AMD’

#### Defining the ‘broad AMD’ phenotype

Past studies that have used UKB data to investigate AMD have employed a broad definition of AMD that considers any participant with an ICD-10 H35.3 diagnosis (‘Degeneration of macula and posterior pole’) as an ‘AMD case’.^16,32^ This diagnostic code is enriched for true AMD cases (i.e., those with dry AMD and/or wet AMD) though is not specific to AMD, as several non-AMD diagnoses also fall under the H35.3 parent code (e.g., angioid streaks of macula; macular cyst, hole, or pseudohole). However, the UKB phenotype data accessible through the NHS and through self-report measures are not sufficiently specific to define a set of AMD-specific cases. For instance, UKB researchers do not have access to ICD-10 codes with greater than 3-digit specificity, meaning it is not possible to determine which participants with a H35.3 code specifically have AMD as encoded by H35.31 (dry/nonexudative AMD) or H35.32 (wet/exudative AMD). Previous studies demonstrate that this approach of using an ICD-10 H35.3 diagnosis as a broadly defined AMD phenotype is able to successfully identify associations that are relevant for true AMD,^16,32^ and results from our own analyses described in this paper also support the relevance and usefulness of using this broadly defined AMD phenotype. We leveraged all relevant sources of disease data to build this broadly defined AMD case set, including as a case any individual with a code or phenotype mapping to ICD-10 H35.3 based on data accessible through NHS sources (in-patient hospitalization records, cancer registry, death records), self-report, and GP records. Throughout the remainder of this paper, we refer to this ICD-10 H35.3 broad definition of AMD as ‘broadly defined AMD’.

### Testing gene-based rare variant associations with broadly defined AMD

We defined high-confidence predicted loss-of-function variants (HC pLOFs) as variants with 1) a VEP^33^ (v109)-predicted consequence of stop gained, frameshift, splice acceptor or splice donor, and 2) designated by LOFTEE^34^ (v104) as a ‘HC’ pLOF. For each gene in the *CFH* region (namely, *CFH* and the five *CFHR*s), we aggregated rare (MAF<1%) HC pLOFs on their own and combined with rare missense variants predicted to be damaging based on different variant effect predictors. More specifically, for each gene we generated multiple rare variant sets based on the following masks: 1) HC pLOFs only; 2) HC pLOFs + missense variants with Combined Annotation-Dependent Depletion PHRED-scaled score (CADD^35^) ≥25; 3) HC pLOFs + missense variants with Rare Exome Variant Ensemble Learner score (REVEL^36^) ≥0.773; 4) HC pLOFs + missense variants with AlphaMissense^37^ score ≥0.564; 5) HC pLOFs + missense variants with ESM1b^38^ ≤-9.39; and 6) HC pLOFs + missense variants with ESM1b ≤-11.52. We selected deleteriousness thresholds for CADD, REVEL and AlphaMissense based on published recommendations,^37,39^ and identified thresholds for ESM1b based on in-house benchmarking. Predicted damaging missense variants are expected to be enriched for variants that result in gene loss-of-function and therefore aggregating them together with HC pLOFs has the potential to increase power for rare variant tests, particularly when pLOF carriers are few.

Gene-based rare variant association testing was performed with Regenie.^40^ For each variant set, we performed burden testing using Regenie’s default burden mask setting (*--build-mask ‘max’*). All analyses adjusted for age at enrollment (‘age’), age^2^, sex, age*sex, age^2^*sex, recruitment region (England, Scotland, Wales), whether the participant was in the general practitioner (GP) dataset, top 10 genetic PCs based on common variants (MAF>1%), top 20 genetic PCs based on rare variants (MAF<1% and MAC≥100), as well as the LOCO (leave-one-chromosome-out) predictions from the Regenie Step 1 model. Genetic input for our Regenie Step 1 models was genome-wide imputed genotypes cleaned and pruned based on the steps described in Backman et al. (2021)^23^ and also restricted to variants with imputation quality score INFO=1. For each gene, we used the aggregated Cauchy association test^41^ (R^42^ package ‘ACAT’) to combine p-values from all burden tests, yielding a single overall p-value for the gene. We refer to these overall p-values for each gene as ‘burden-ACAT p-values’.

We implemented analytical models that adjusted for known independent AMD genetic signals at the *CFH* region as follows: 1) adjustment for *CFH* Y402H (rs1061170) and *ΔCFHR3-CFHR1* (using rs6677604 as a proxy for the deletion based on nearly perfect LD), and 2) adjustment for *CFH* Y402H, Δ*CFHR3-CFHR1*, and six of the eight independent AMD-associated variants at the *CFH* region as reported by Fritsche et al. (2016).^14^ One of the eight Fritsche-identified *CFH*-region variants (rs570618) is in near perfect LD with *CFH* Y402H (among individuals with European ancestry) and thus is already controlled for by including Y402H as a covariate. A second Fritsche-identified variant (rs61818925) is a common intergenic SNP missing for 27% of our analytic sample due to lower imputation quality. We decided to not include this SNP as a covariate in our main analyses. However, we performed supplemental sensitivity analyses that adjusted for all variants plus rs61818925, restricting to the 73% of individuals with higher quality genotypes for this SNP.

We present results for the two aforementioned variant-adjustment models, one less rigorously controlled and one more rigorously controlled. The more rigorous adjustment model, which controls for *CFH* Y402H, Δ*CFHR3-CFHR1*, and several additional *CFH*-region AMD-associated variant signals identified by Fritsche et al., is important as a means of decreasing as much as possible the potential confounding influence of LD with other AMD-associated variants at the *CFH*-region. However, this most rigorous adjustment model may at times result in a degree of overcorrection, specifically if the tested rare variants are causal and have LD with the adjusted variants. For this reason, we also present results from a less rigorously adjusted model, though in this model we still adjust for *CFH* Y402H and *ΔCFHR3-CFHR1* as both are established as likely causal variants with high frequencies and large effects on AMD, making it important to condition out their influence in our rare burden analyses.

### Testing gene-based rare variant associations with ‘strictly defined AMD’

#### Defining the ‘strict AMD’ phenotypes

General practitioner (GP) electronic health records were available for approximately half of UKB participants, and these records include diagnostic subcodes specific to AMD, including separate codes for dry AMD and wet AMD. This offers greater phenotypic specificity compared with the broadly defined AMD phenotype. However, a major drawback is that the case counts for these AMD-specific codes, based solely on the available GP data, are quite small with the result that rare variant analyses of these strictly defined AMD phenotypes are expected to be largely underpowered to detect statistically significant associations. Nevertheless, we performed rare burden analyses using these AMD-specific phenotypes to see whether we might identify any evidence for rare variant associations with dry or wet AMD subtypes. We were particularly interested in examining potential associations with dry AMD, considering a lack of effective therapies for this most common AMD subtype.

GP records were extracted from the TPP system which employs Clinical Terms Version 3 (CTV3/Read v3) codes, Read v2 codes, and Local TPP codes. Only CTV3 and Read v2 codes were used for AMD diagnoses. We identified AMD cases as individuals with a code for non-exudative (dry) AMD (F4251 for both CTV3 and Read v2; this code corresponds to ICD-10 H35.31) or exudative (wet) AMD (F4252 for both CTV3 and Read v2; this code corresponds to ICD-10 H35.32). We performed analyses for dry and wet AMD separately, and additionally tested dry and wet AMD cases combined. In the remainder of this paper, we refer to these AMD specific phenotypes as ‘strictly defined AMD’. We defined controls as participants represented in the GP records who were lacking one of these AMD-specific diagnostic codes.

#### Testing gene-based rare variant associations with strictly defined AMD

Using Regenie, we tested gene-based rare variant burden associations with these three strictly defined AMD groupings. Analyses mirrored those for broadly defined AMD, controlling for the same covariates except for the GP indicator variable (since these analyses were restricted to participants in the GP dataset), and also controlling for different numbers of known *CFH*-region AMD-associated signals.

#### Testing gene-based rare variant associations with AMD-predictive retinal thickness measures Quality control of OCT data

About 16% of the UKB cohort underwent retinal optical coherence tomography (OCT), which involves a 3-dimensional scan and photograph of the retina as well as a magnified photograph of the fundus. Investigators subsequently applied Topcon Advanced Boundary Segmentation (TABS) software to these imaging data to derive quantitative measures of various retinal layers and other ocular elements.^19,20^ These quantitative OCT measures were later shared with the UKB study and made available to all UKB-approved researchers. In total, 40 different OCT retinal layer thickness measures have been generated separately for the right and left eyes of participants, yielding 80 OCT layer thickness data fields (UKB Category 100079).

As recommended by the UKB study and prior studies that have analyzed these OCT data, we applied quality control filters to remove the subset of participants with OCT measures derived from low-quality imaging. There were 82,848 participants with OCT QC metrics for the left eye (67,275 from Instance 0 [initial assessment visit] and 15,573 from Instance 1 [first repeat assessment visit]) and 82,839 participants with OCT QC metrics for the right eye (67,278 from Instance 0 and 15,561 from Instance 1), including 82,833 participants with QC metrics for both left and right eyes (these quality control measures are provided as UKB Category 100116). There was no sample overlap for OCT data from Instances 0 and 1 (i.e., participants underwent OCT imaging at a single visit only). We applied OCT QC as described in Ko et al. (2017)^19^ and Patel et al. (2016).^20^ This involved excluding left or right eyes with image quality score < 45 as well as those in the poorest 20% for any of the other segmentation indicators: ILM indicator, macula center aline, macular center frame, max motion delta, max motion factor, min motion correlation, and valid count. We observed that Instance 1 metrics seemed to reflect lower average image and segmentation quality for the OCT data generated at Instance 1 as compared with Instance 0, so we identified the ‘poorest 20%’ segmentation quality thresholds based on Instance 0 only, then applied these same thresholds to both Instance 0 and Instance 1. The ‘poorest 20%’ segmentation quality thresholds for the left eye were: ILM <7363.592, validity count <870, min motion correlation <0.3290116, max motion delta >5.41211, and max motion factor >5.09961. Applying these thresholds and also excluding eyes with image quality score <45 yielded 47,198 left eyes (40,664 from Instance 0 and 6,534 from Instance 1) passing QC. The ‘poorest 20%’ segmentation quality thresholds for the right eye were: ILM <6643.62, validity count <886, min motion correlation <0.2946708, max motion delta >5.56055, and max motion factor >5.27539. Applying these thresholds and also excluding eyes with image quality score <45 yielded 44,712 right eyes (40,122 from Instance 0 and 4,590 from Instance 1) passing QC. A total of 32,042 participants had both left and right eyes that passed strict QC, and for these individuals we averaged left and right eye values for each of the 40 different OCT layer thickness measures for use in our OCT analyses. For the remaining individuals we used values derived from the single eye that passed QC. This left nearly 60,000 participants with good quality OCT measures (before restricting to European ancestry).

A prior analysis of OCT retinal layer thickness measure associations with broadly defined AMD using data from the UKB study, performed by Zekavat et al.^16^, applied more lenient QC to the OCT data (excluding eyes with: image quality score <40, those in the poorest 10% of the ILM indicator, and those with OCT measure values >2.5 standard deviations from the mean). We tested application of these lenient QC filters and found that our ‘strictly defined AMD’ phenotype significantly and more strongly associated with OCT measures processed using the Ko et al. (2017)^19^ and Patel et al. (2016)^20^ approach (we detail these associations in the Results section) as compared with OCT measures processed using the more lenient QC, despite the lenient QC leaving 21% more participants for analysis (AMD associations with OCT data processed using lenient QC are available in *Table S14*). Application of the Ko et al. (2017) and Patel et al. (2016) QC approach therefore yielded OCT phenotypes that were stronger predictors of AMD, and for this reason we moved forward with this QC approach.

#### Testing OCT phenotype associations with prevalent and incident AMD

We tested the 40 OCT retinal layer thickness phenotypes for association with AMD, prioritizing associations with the AMD-specific strict AMD phenotype but also testing associations with broadly defined AMD, and we examined both prevalent and incident AMD. Prevalent AMD was defined as having the relevant (i.e., strict or broad) AMD diagnosis at any point in time, while for incident AMD we excluded as cases individuals with a first occurrence of AMD diagnosis prior to or the same date as their UKB OCT assessment. Though Zekavat et al.^16^ previously analyzed OCT measure associations with prevalent and incident AMD using UKB data, our analyses have a few notable differences: 1) Zekavat et al. focused specifically on broadly defined AMD, whereas we also test associations with strictly defined AMD; 2) Zekavat et al. combined certain OCT thickness measures through summation and analyzed the resulting combined phenotypes (e.g., summing thicknesses for the photoreceptor layer inner segment [ELM-ISOS] and the photoreceptor layer outer segment [ISOS-RPE] and analyzing the resultant phenotype as a measure of the overall photoreceptor), whereas we analyze each measure separately; and 3) we implemented a stricter QC approach for the OCT data.

We applied inverse normal transformation to each OCT phenotype and used linear regression to test association of OCT measure and AMD, regressing the OCT measure onto AMD and adjusting for age at OCT imaging (‘age _OCT_’), age_OCT_ ^2^, sex, age_OCT_ *sex, age ^2^*sex, OCT assessment instance, GP status (only included when analyzing broadly defined AMD), and country of recruitment. Though we expect certain retinal changes to precede AMD, we modeled AMD status as a predictor variable to facilitate interpretation of how cases and controls compare with respect to OCT measures. These analyses were restricted to unrelated participants, and were performed using R.^42^

#### Testing gene-based rare variant associations with AMD-associated OCT phenotypes

We next examined gene-based rare variant associations with the specific OCT phenotypes that we identified as strongly and significantly predictive of AMD (see Results). As with the rare variant analyses of broadly defined AMD, we performed burden tests of rare HC pLOFs aggregated on their own and in combination with predicted-damaging missense variants for association with the top OCT phenotypes. For each gene, burden p-values for all variant sets were aggregated using ACAT to yield a single ‘burden-ACAT p-value’ per gene. Analyses adjusted for age_OCT_, age_OCT_ ^2^, sex, age_OCT_ *sex, age_OCT_ ^2^*sex, OCT assessment instance, 10 PCs based on common variants, 20 PCs based on rare variants, and Regenie Step 1 predictions. We also adjusted for different numbers of known independent AMD signals at the *CFH* region, as described previously.

Our primary analyses excluded all AMD cases (broadly and strictly defined) as a means of examining gene-based rare variant burden associations with AMD-related phenotypes in a manner that is semi-orthogonal to the AMD case-control analyses. Such analyses can also offer additional evidence regarding whether OCT thickness changes may be more likely to precede or follow AMD diagnosis. We also performed supplemental analyses that retained AMD cases.

#### Examining possible interaction between *CFHR5* rare variant burden and *CFH Y402H*

Following identification of the association of *CFHR5* rare predicted-damaging variants with AMD protection (see Results section), we examined whether the *CFHR5* protective association may be greater for individuals carrying a *CFH* Y402H risk allele. We used Regenie to perform formal tests of interaction between *CFH* Y402H and *CFHR5* rare variant burden, modeling the outcome as the broadly defined AMD phenotype to maximize analytical power for detecting AMD associations. We carried out similar interaction analyses with OCT phenotype as the outcome.

#### Testing gene-based rare variant associations with serum protein levels of *CFH*-region genes

Approximately 10% of the UKB participants have serum protein levels assayed for a subset of human proteins (nearly 3,000 proteins) using the Olink Explore panel. The assayed proteins include *CFH, CFHR2, CFHR4* and *CFHR5*, and do not include *CFHR1* or *CFHR3*. We leveraged these proteomics data to examine the protein-specificity of each gene-based rare variant set used in our AMD and OCT phenotype analyses (e.g., to what extent do *CFH* rare variant sets associate with *CFH* serum protein versus serum protein of the *CFHR*s?).

We accessed protein measurements that were log_2_ transformed and that had already undergone intra- and inter-individual normalization as detailed in Sun et al. (2023).^21^ Prior to analyses we applied rank-based inverse normalization to each protein. Using burden tests, we examined each gene-based rare variant set for association with serum protein levels of *CFH, CFHR2, CFHR4* and *CFHR5*. Covariates adjusted for included age, age^2^, sex, age*sex, age^2^*sex, top 10 genetic PCs based on common variants, top 20 genetic PCs based on rare variants, Olink sample batch, and different numbers of known independent AMD variant signals at the *CFH* region.

## RESULTS

### Rare predicted-damaging variants in CFHR5 and CFH significantly associate with AMD

For each gene at the *CFH* locus, we performed gene-based rare (MAF<1%) variant burden analyses of predicted-damaging variants to help clarify the gene’s potential role in AMD. We used several different functionally defined variant masks, and we implemented both a more lenient adjustment model controlling for *CFH* Y402H and *ΔCFHR3-CFHR1* and a more rigorous adjustment model controlling for these and several additional AMD-associated variants at the *CFH* region. These analyses employed a broad definition of AMD that considers any participant with an ICD-10 H35.3 diagnosis (‘Degeneration of macula and posterior pole’) as an ‘AMD case’ (see Methods section for further details). Analyses included 10,700 broadly defined AMD cases and 396,099 controls.

We initially aggregated rare burden results for each gene (separately for each adjustment model) using the ACAT approach, to reduce multiple testing burden and enable identification of AMD-associated genes. Setting a strict Bonferroni-corrected significance threshold of p < 0.05/12 = 0.0041 (6 genes * 2 adjustment models), we observe significant burden-ACAT p-values for *CFH* and *CFHR5* (**Table 1**). *CFH* burden-ACAT p-values are strongly significant across both variant adjustment models. *CFHR5* yields a significant burden-ACAT p-value value when adjusting for *CFH* Y402H and *ΔCFHR3-CFHR1* (p=1.6×10^−3^) and a suggestive result when adjusting for several additional *CFH*-region AMD signals (p=0.02). None of the other *CFHR* genes yield significant burden-ACAT p-values in either adjustment model, though carrier counts are low and statistical power is likely limited, particularly for *CFHR1, CFHR2* and *CFHR3*. For *CFHR2* there is a nominal result (p=0.04), but examination of the individual variant set results (**Table S1**) reveals this is driven by *CFHR2* variant sets with particularly few carriers and is unlikely to be a reliable result.

**Table 1.** Gene-based rare (MAF<1%) variant burden associations with broadly defined AMD.

| Gene | Adjusting for Y402H and $\Delta CFHR3-CFHR1$ | | | Adjusting for Y402H and $\Delta CFHR3-CFHR1$, plus six variants | | |
| --- | --- | --- | --- | --- | --- | --- |
|  | Total samples (Cases) | Total carriers (Case carriers) | Burden-ACAT P-value | Total samples (Cases) | Total carriers (Case carriers) | Burden-ACAT P-value |
| *CFH* | 406,799 (10,700) | 284 (35) | $2.8 \times 10^{-14}$ | 397,241 (10,409) | 278 (34) | $8.3 \times 10^{-14}$ |
| *CFHR5* | 406,799 (10,700) | 7,860 (156) | $1.6 \times 10^{-3}$ | 397,241 (10,409) | 7,727 (154) | 0.02 |
| *CFHR2* | 406,799 (10,700) | 681 (15) | 0.04 | 397,241 (10,409) | 674 (15) | 0.04 |
| *CFHR1* | 390,187 (10,376) | 195 (6) | 0.18 | 380,847 (10,085) | 193 (6) | 0.16 |
| *CFHR3* | 391,521 (10,411) | 434 (16) | 0.30 | 382,079 (10,120) | 428 (16) | 0.25 |
| *CFHR4* | 406,786 (10,700) | 1,596 (43) | 0.96 | 397,230 (10,409) | 1,573 (41) | 0.97 |

For each gene, p-values from multiple rare (MAF<1%) variant masks were combined using the aggregated Cauchy association test (ACAT), yielding a single overall ‘burden-ACAT p-value’. Results are presented for two variant-adjustment models. *CFH* Y402H (rs1061170) was controlled for directly. $\Delta CFHR3-CFHR1$ was controlled using rs6677604 based on near-perfect LD. ‘Plus six variants’ refers to six of the independently AMD-associated *CFH*-region variants identified by Fritsche et al. (2016)<sup>14</sup>: rs191281603, rs187328863, rs148553336, rs10922109, rs121913059, rs35292876. We set a strict Bonferroni-corrected significance threshold of $p < 0.0041$ .

### CFHR5 rare pLOF and predicted-damaging missense variants associate with decreased AMD risk

We followed up on the significant burden-ACAT results by exploring individual variant set burden results for *CFH* and *CFHR5*. We observed four unique variant masks each for *CFH* and *CFHR5*, as some variant masks contained identical variants. We excluded these duplicate masks and then examined the *CFH* and *CFHR5* rare burden results, setting a strict Bonferroni-corrected significance threshold of p < 0.05/16 = 0.0031 (2 genes * 4 variant masks * 2 adjustment models).

Considering extensive existing genetic evidence for the role of *CFH* in AMD, our most novel finding is that rare *CFHR5* pLOF variants independently associate with protection from AMD. The strongest result is observed for *CFHR5* pLOFs combined with AlphaMissense-predicted-damaging missense variants, which significantly associate with a 25% decreased odds of AMD (p=7.1×10^−4^) when adjusting for Y402H and *ΔCFHR3-CFHR1*, and this association remains suggestive in the more rigorous model adjusted for additional variants (p=0.009). Aggregated *CFHR5* pLOFs tested on their own also yield an AMD-protective association that is significant in the model adjusted for Y402H and *ΔCFHR3-CFHR1* (p=0.0017) and suggestive in the more rigorous model (p=0.019) (**Figure 1**; **Table 2**).

**Table 2.** *CFHR5* and *CFH* individual rare (MAF<1%) variant set burden associations with broadly defined AMD.

| | | Adjusting for Y402H and $\Delta CFHR3-CFHR1$ | | | | Adjusting for Y402H and $\Delta CFHR3-CFHR1$, plus six variants | | | |
| --- | --- | --- | --- | --- | --- | --- | --- | --- | --- |
| Gene | Variant Set | Total samples (Cases) | Total carriers (Case carriers) | OR | P-value | Total samples (Cases) | Total carriers (Case carriers) | OR | P-value |
| *CFHR5* | HC pLOF + AM≥0.56 | 406,799 (10,700) | 6,987 (133) | 0.75 | $7.1 \times 10^{-4}$ | 397,241 (10,409) | 6,871 (132) | 0.80 | 0.009 |
|  | HC pLOF |  | 6,354 (122) | 0.76 | 0.0017 |  | 6,249 (121) | 0.81 | 0.019 |
| | HC pLOF + ESM1b≤-11.52 | | 6,768 (132) | 0.77 | 0.0024 | | 6,654 (131) | 0.82 | 0.027 |
| | HC pLOF + ESM1b≤-9.39 | | 7,860 (156) | 0.80 | 0.0042 | | 7,727 (154) | 0.84 | 0.035 |
| *CFH* | HC pLOF + AM≥0.56 | 406,799 (10,700) | 284 (35) | 5.95 | $4.6 \times 10^{-15}$ | 397,241 (10,409) | 278 (34) | 5.90 | $1.4 \times 10^{-14}$ |
| | HC pLOF | | 81 (11) | 7.11 | $1.6 \times 10^{-6}$ | | 81 (11) | 6.95 | $2.0 \times 10^{-6}$ |
| | HC pLOF + CADD≥25 | | 170 (22) | 6.54 | $7.8 \times 10^{-11}$ | | 166 (21) | 6.24 | $4.5 \times 10^{-10}$ |
| | HC pLOF + REVEL≥0.77 | | 136 (15) | 5.59 | $4.1 \times 10^{-7}$ | | 134 (15) | 5.60 | $4.0 \times 10^{-7}$ |

Variant set 'HC pLOF' tested aggregated rare (MAF<1%) HC pLOF variants on their own, and other variant sets combined HC pLOFs with missense variants predicted as damaging based on AlphaMissense score ≥ 0.56, ESM1b ≤ -11.52, ESM1b ≤ -9.39, CADD ≥ 25, or REVEL ≥ 0.77. For each gene, only variant masks representing unique variant sets are shown. Results are presented for two variant-adjustment models. *CFH* Y402H (rs1061170) was controlled for directly. $\Delta CFHR3-CFHR1$ was controlled using rs6677604 based on near-perfect LD. 'Plus six variants' refers to six of the independently AMD-associated *CFH*-region variants identified by Fritsche et al. (2016):<sup>14</sup> rs191281603, rs187328863, rs148553336, rs10922109, rs121913059, rs35292876. We set a strict Bonferroni-corrected significance threshold of $p < 0.0031$ , and $p < 0.05$ was considered suggestive. 'OR' is odds ratio.

**Figure 1.**
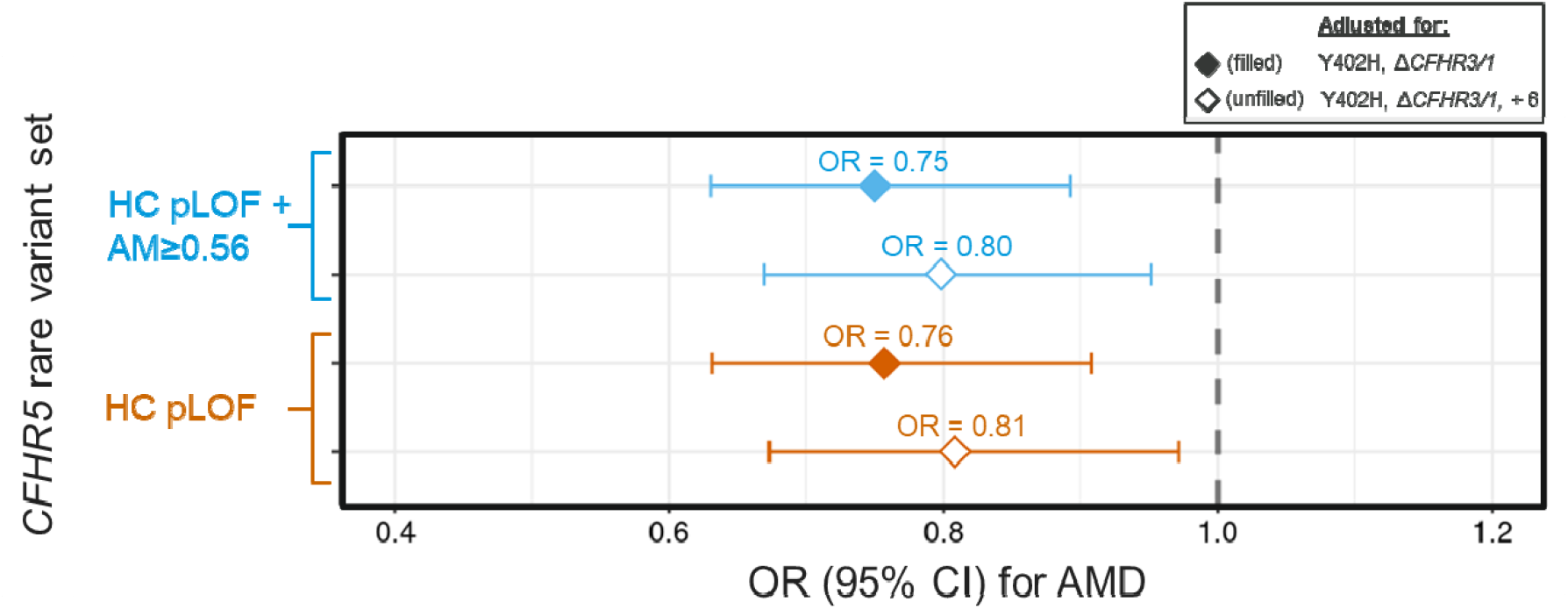
Rare *CFHR5* pLOFs and predicted-damaging missense variants significantly associate with protection from AMD. Shown are associations of broadly defined AMD with CFHR5 rare (MAF<1%) HC pLOFs aggregated on their own (‘HC pLOF’) and in combination with AlphaMissense-predicted damaging missense variants (‘HC pLOF + AM≥0.56’), both for the model adjusting for CFH Y402H and *ΔCFHR3-CFHR1* (‘*ΔCFHR3/1’*) and for the more rigorous model that adjusts for an additional six independently AMD-associated CFH-region variants. ‘OR’ is odds ratio. ‘CI’ is confidence interval.

For *CFH*, pLOF and predicted-damaging missense variants occurred at very low frequency (MAF<0.01% for all *CFH* rare variants), perhaps reflecting selection against CFH-damaging variants. Despite very low *CFH* rare variant carrier counts, we see across both adjustment models that pLOFs aggregated on their own and in combination with predicted-damaging missense variants very strongly and significantly associate with increased risk of AMD (Table 2). *CFH* pLOFs combined with AlphaMissense-predicted-damaging missense variants yield the strongest result, associating with a nearly 6-fold greater odds of broadly defined AMD in the more rigorous adjustment model (p=1.4×10). These results are consistent with *CFH* and *CFHR5* having opposing effects on AMD, with loss of function expected to increase AMD risk for *CFH* and protect against AMD for *CFHR5*.

In supplemental sensitivity analyses that additionally control for the *CFH*-region SNP rs61818925, identified as independently associated with AMD by Fritsche et al^14^. but excluded from our main analyses due to being missing for a large portion of our analytic sample, the association of *CFHR5* rare predicted-damaging variants with AMD protection remains suggestive (p=0.037) despite a 27% decrease in analytic sample size (Table S1). The association of *CFH* rare burden with increased AMD risk also remains.

As detailed in **Appendix A**, we used Olink-based proteomics data (available for 44,088 participants in our analytic sample) to validate all *CFHR5* and *CFH* rare variant sets as strongly and significantly associating with reductions in *CFHR5* and *CFH* circulating protein levels, respectively; and as displaying high specificity for their encoded proteins, with greatest specificity observed when implementing the more rigorous variant adjustment model that conditions on *CFH* Y402H, *ΔCFHR3-CFHR1*, and several other AMD-associated variants in the *CFH* region (**Table S2**).

### Two pLOF variants explain much of the *CFHR5* rare burden association with AMD protection

In follow-up analyses, we observed that the *CFHR5* rare variant burden association with protection from broadly defined AMD is largely driven by rare variants with higher relative frequencies (0.1%<MAF<1%; **Table S3**). Our set of rare *CFHR5* pLOFs combined with AlphaMissense-predicted-damaging missense variants, which yields the strongest AMD-protective association, includes three such variants: two pLOF frameshift variants and one missense variant. The two pLOF variants are different alternate alleles for the same multiallelic locus: rs565457964-CA (HGVSp: p.Glu163ArgfsTer35) and rs565457964-CAA (HGVSp: p.Glu163LysfsTer10). rs565457964-CA has an alternate allele frequency (AAF) of 0.45% in our analytic sample (restricted to individuals of European genetic ancestry), and rs565457964-CAA has AAF=0.27%. The missense variant is rs41299613-C (HGVSp: p.Cys208Arg) with AAF=0.32%, and is predicted as damaging based on AlphaMissense score = 0.93. Further examination reveals that the missense variant rs41299613-C occurs almost exclusively in combination with the pLOF rs565457964-CA (D’=0.98, r^2^=0.68), with conditional analyses demonstrating that rs565457964-CA can fully account for an association of the missense variant with AMD (**Table S4**).

In individual variant tests of association with AMD adjusting for Y402H and *ΔCFHR3-CFHR1*, we observe AMD-protective associations for both *CFHR5* pLOFs (**Table S5**). A follow-up investigation into nearby coding variants in LD with either of the two pLOFs revealed no variants that could better account for the AMD association, further supporting *CFHR5* genetic inhibition as the causal mechanism underlying the observed AMD protection (**Appendix B**).

The *CFHR5* pLOF rs565457964-CAA has approximately 10-fold higher frequency in the Finnish genetic ancestry group as compared with the non-Finnish European ancestral group.^43^ A test of this variant for association with AMD would therefore have much greater power in a large Finnish ancestry sample, though it may be more susceptible to LD-induced confounding due to much higher variant frequency. The latest public release (r11) of results from the FinnGen Biobank study^44,45^ indeed shows strongly significant associations of rs565457964-CAA with protection from dry or wet AMD (OR=0.51, p=4.6×10^−54^), dry AMD (OR=0.49, p=2.2×10^−42^) and wet AMD (OR=0.48, p=1.6×10^−34^), when not adjusting for other known *CFH*-region AMD variant signals (**Table S6**). In addition, conditional analyses by FinnGen show that the association of rs565457964-CAA with AMD protection remains significant (OR=0.78, p=3.1×10^−8^) after controlling for certain SNPs that tag several of the *CFH*-region AMD-associated variants controlled for in our analyses, including a SNP in perfect LD (r^2^=1) with *CFH* Y402H and SNPs that partially tag *ΔCFHR3-CFHR1* (**Table S7**). FinnGen fine-mapping of the AMD GWAS signals (using SuSIE^46^) shows that rs565457964-CAA is the sole coding variant in a 95% credible set that includes 10 SNPs in high LD with one another. The SNPs in this credible set exhibit Finnish-ancestry-based LD patterns with our nine *CFH*-region control variants that are similar to the LD between rs565457964-CAA and the control variants as estimated using our non-Finnish European ancestry sample (**Table S8**). This suggests that the FinnGen rs565457964-CAA association with AMD protection is likely to remain after adjusting for *CFH* Y402H, *ΔCFHR3-CFHR1*, and several additional *CFH*-region AMD signals, as it does in our UKB rare variant analyses (**Table 2**; **Table S5**).

FinnGen also reports results for *CFHR5* HC pLOF rs565457964-CA, which is very rare among those with Finnish genetic ancestry (MAF=0.08%) and therefore likely less susceptible to LD-induced confounding as compared with rs565457964-CAA. They find that this ultra-rare variant nominally associates with decreased risk for dry or wet AMD (OR=0.33, p=0.002), wet AMD (OR=0.41, p=0.011), and dry AMD (OR=0.54, p=0.048) (**Table S6**). The FinnGen study findings for *CFHR5* pLOFs rs565457964-CAA and rs565457964-CA lend important additional support to our UKB-based observations that *CFHR5* rare pLOFs and other predicted-damaging variants independently associate with decreased AMD risk. In combination, these results from UKB and FinnGen analyses strongly support *CFHR5* genetic inhibition as protective against AMD, including both dry and wet forms.

In addition, FinnGen reports associations for a phenotype matching our ‘broadly defined AMD’, with results exhibiting consistency with the AMD-specific associations reported by the FinnGen study (**Table S6**). This observed consistency supports use of the ‘broad AMD’ phenotype as a means of gaining insight into associations with AMD-specific phenotypes.^44,45,47^

### *CFHR5* rare variant association with AMD protection appears stronger in *CFH* Y402H risk allele carriers

We examined whether the association of *CFHR5* rare predicted-damaging variants with AMD protection differed by *CFH* Y402H genotype. **Figure 2** presents estimated associations between broadly defined AMD and *CFHR5* rare pLOFs combined with AlphaMissense-predicted-damaging missense variants, stratified by number of *CFH* Y402H risk allele copies and adjusted for *ΔCFHR3-CFHR1*. We see that the estimated associations of *CFHR5* rare variant burden with AMD are more strongly protective for *CFH* Y402H risk allele carriers.

**Figure 2.**
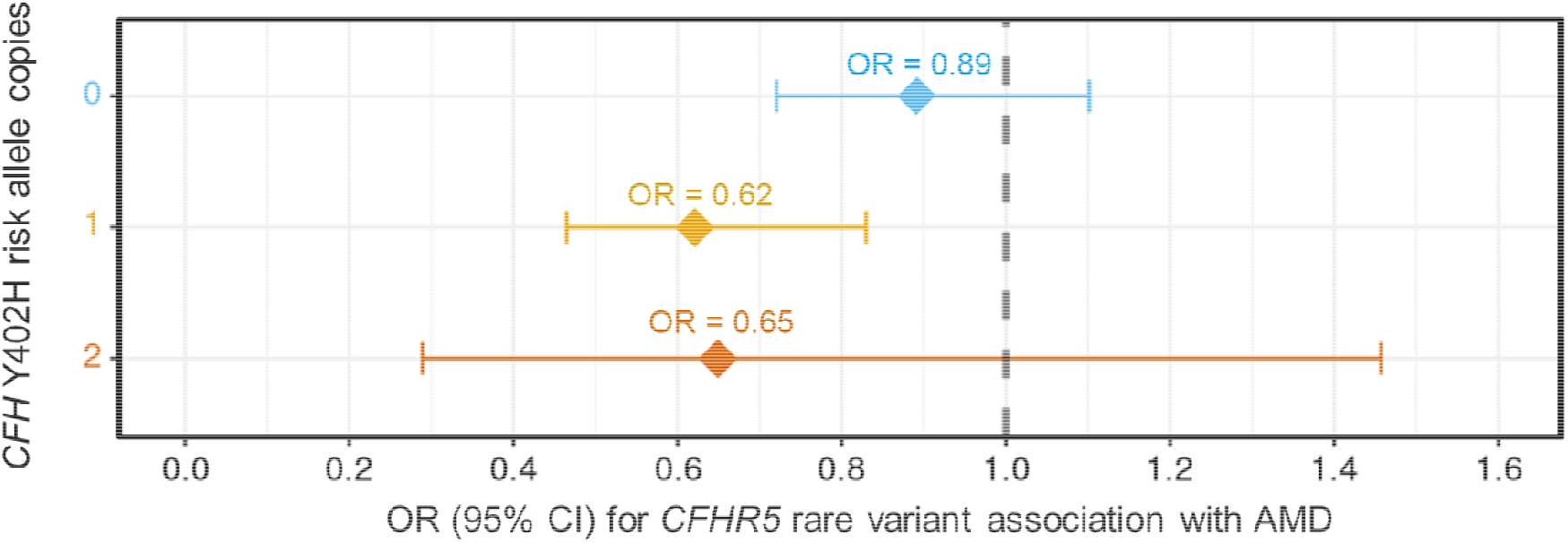
*CFHR5* rare predicted-damaging variant associations with broadly defined AMD appear stronger among *CFH* Y402H risk allele carriers. We estimated associations of broadly defined AMD with *CFHR5* rare (MAF<1%) HC pLOFs combined with AlphaMissense-predicted-damaging missense variants, stratified by number of *CFH* Y402H (rs1061170-C) copies (y-axis). Number of AMD cases and *CFHR5* rare variant carriers for each Y402H genotype are provided in **Table S9**. These analyses adjusted for *ΔCFHR3-CFHR1* (using tag SNP rs6677604). Y402H carriers display estimates consistent with stronger protection from *CFHR5* rare variants. ‘OR’ is odds ratio. ‘CI’ is confidence interval.

In formal tests for interaction, we observe interaction p=0.05 in the model adjusting for Y402H and *ΔCFHR3-CFHR1*, and interaction p=0.04 for the more rigorously adjusted model that controls for several additional *CFH*-region AMD-associated variants (**Table S10**). These results are consistent with the possibility of a greater AMD protective benefit conferred by *CFHR5* rare variants for the CFH Y402H risk allele carriers.

### *CFH* pLOF and predicted-damaging missense variants associate with increased risk for dry AMD

We used participants with GP data (n=189,515), which includes more fine-grained diagnostic codes, to test for associations with dry AMD (n=253) and wet AMD (n=112) subtypes. We tested *CFHR5* and *CFH* rare (MAF<1%) variant burden for association with each subtype separately, as well as with dry and wet AMD combined. For these analyses of strictly defined AMD, and for the remaining *CFHR5* and *CFH* rare variant analyses described in the Results section, we used only two variant masks for each gene: 1) HC pLOFs combined with AlphaMissense-predicted-damaging missense variants, and 2) HC pLOFs on their own. The former variant mask offers greater power for rare burden analyses and yields the strongest associations with broadly defined AMD for both *CFHR5* and *CFH*; the latter variant mask offers the clearest interpretation of functional consequence by virtue of only including pLOFs.

For *CFHR5*, low statistical power prevents detection of an expected protective association with the strict AMD phenotypes (Table S11). In contrast, both *CFH* rare variant sets significantly associate with increased risk of strictly defined AMD, with *CFH* pLOFs combined with AlphaMissense-predicted-damaging variants yielding a 20-fold increased odds of dry or wet AMD (OR=20.3, p=4.4×10^−6^) and a 27-fold increased odds of dry AMD specifically (OR=26.8, p=7.0×10^−6^) for the more rigorously adjusted model (**Table S11**). These findings add support to evidence linking the *CFH* region with dry AMD.^14^ Prior studies have also linked the *CFH* region with wet AMD,^14^ though our analyses were underpowered to detect associations with wet AMD specifically. We note that the AMD-specific risk estimates for *CFH* are several times larger than the corresponding estimates for broadly defined AMD (see **Table 2, Table S1**), and that combined with FinnGen results showing larger association estimates for strict versus broad AMD (**Table S6**), this supports the possibility that true AMD-specific associations with rare, damaging variants may be greater in magnitude than what we estimate using the broadly defined AMD phenotype.

### Changes in thickness of the photoreceptor layer of the retina precede AMD diagnosis

In addition to our AMD case-control analyses, we sought to examine evidence from complementary rare variant analyses of AMD-predictive quantitative retinal phenotypes. To do this, we first tested the association of 40 OCT retinal layer thickness measures with AMD, to identify those measures most strongly predictive of AMD. As the phenotype most relevant to AMD, we prioritized OCT associations with strictly defined AMD, and we tested dry and wet AMD both combined and separately. We also tested OCT associations with broadly defined AMD and examined consistency of results with those from tests of strictly defined AMD. Our primary analyses focused on prevalent AMD to maximize power, but we also performed supplemental analyses of incident AMD (defined as having a first occurrence of AMD diagnosis after the OCT assessment date).

Our analyses of prevalent AMD associations with OCT phenotypes included 160 tests (40 OCT measures * 4 AMD phenotypes). Complete results are provided in **Table S12**. We set a strict Bonferroni-corrected significance threshold of 0.05/160 = 3.1×10^−4^. As shown in **Table 3**, for strictly defined AMD, we observe those with a dry or wet AMD diagnosis to have significantly thicker inner segment of the photoceptor layer (labeled by the UKB study as ‘ELM-ISOS’ thickness), particularly when measured at the inner subfield (+0.91 SD thickness for AMD cases, p=1.3×10^−6^); significantly thinner outer segment of the photoreceptor layer (labeled by the UKB study as ‘ISOS-RPE’ thickness), particularly when measured at the central subfield (-0.79 SD, p=3.4×10^−5^); and significantly decreased macular thickness as measured at the inner inferior subfield (-0.84 SD, p=1.7×10^−5^). In supplemental analyses restricting to incident cases, these same OCT measures maintained strong associations (**Table S13**). Tests of OCT phenotype associations with dry and wet AMD separately were generally underpowered due to small case counts, limiting ability to examine whether these AMD subtypes differentially associate with certain OCT measures. However, we observe that top associations for each subtype are largely consistent with the OCT phenotypes most strongly associated with combined dry or wet AMD (**Table S12; Table S13)**.

**Table 3.** OCT phenotypes significantly associating with strictly or broadly defined AMD.

|  | Strict AMD (Dry or Wet) |  |  | Broad AMD (ICD-10 H35.3) |  |  |
| --- | --- | --- | --- | --- | --- | --- |
| OCT phenotype | Total samples (Cases) | Beta | P-value | Total samples (Cases) | Beta | P-value |
| Thickness of the inner segment of the photoreceptor layer (inner subfield) | 18,055 (27) | 0.91 | $1.3 \times 10^{-6}$ | 43,379 (942) | 0.21 | $2.3 \times 10^{-10}$ |
| Macular thickness (inner inferior subfield) | 15,853 (25) | -0.84 | $1.7 \times 10^{-5}$ | 37,114 (798) | -0.14 | $1.1 \times 10^{-4}$ |
| Thickness of the outer segment of the photoreceptor layer (central subfield) | 18,116 (27) | -0.79 | $3.4 \times 10^{-5}$ | 43,530 (947) | -0.24 | $2.9 \times 10^{-13}$ |
| Macular thickness (inner nasal subfield) | 15,905 (25) | -0.63 | 0.001 | 37,231 (801) | -0.14 | $5.9 \times 10^{-5}$ |
| Full photoreceptor layer thickness (central subfield) | 18,116 (27) | -0.59 | 0.002 | 43,530 (947) | -0.13 | $8.2 \times 10^{-5}$ |
| Average ganglion cell inner plexiform layer thickness | 18,055 (27) | -0.45 | 0.01 | 43,379 (942) | -0.12 | $4.0 \times 10^{-5}$ |
| Retinal pigment epithelium thickness (central subfield) | 10,702 (11) | 0.44 | 0.14 | 24,769 (424) | 0.21 | $1.8 \times 10^{-5}$ |

Tests of OCT phenotype associations with broadly defined AMD show that the top OCT predictors of broad AMD are the same as those identified for strictly defined AMD (**Table 3; Table S12**). Broadly defined AMD most strongly associates with thinner outer segment of the photoreceptor layer, particularly as measured at the central subfield (-0.24 SD, p=2.9×10^−13^); and thicker inner segment of the photoreceptor layer, particularly at the inner subfield (+0.21 SD, p=2.3×10^−10^). Results were similar when restricting to incident cases of broadly defined AMD (**Table S13**). The consistency of these top OCT measure associations for broadly defined AMD with those observed for strictly defined AMD further supports the relevance of the broadly defined AMD phenotype to AMD-specific diagnoses.

Sensitivity analyses testing OCT phenotype associations with AMD and additionally controlling for smoking and body mass index (BMI), which are known risk factors for AMD, yield no meaningful difference in results (**Table S12; Table S13**).

As the photoreceptor layer outer and inner segments are directly adjacent to one another, we wondered whether the observed significant findings for both segments may reflect correlation between rather than independent associations with AMD. As detailed in **Appendix C**, conditional analyses demonstrate that each segment has an association with AMD that is independent of the other segment.

### CFHR5 and CFH rare variants associate with photoreceptor layer thickness independently of AMD

We tested *CFHR5* and *CFH* rare variant burden associations with outer segment of the photoreceptor layer (measured at the central subfield), inner segment of the photoreceptor layer (measured at the inner subfield), and macular thickness at the inner inferior subfield, which were the OCT phenotypes most strongly predictive of strictly and broadly defined AMD. We report results from analyses that excluded all AMD cases (strictly or broadly defined), as a means of investigating the roles of *CFHR5* and *CFH* in AMD using an approach that is semi-orthogonal to our AMD case-control analyses.

For *CFHR5*, in addition to testing MAF<1% variant sets, we tested MAF<0.1% variant sets to examine whether ultra-rare variants may also contribute to an association (i.e., rare variants other than the higher frequency rare variants observed to be driving the *CFHR5* association with protection from AMD). Such ultra-rare variants are typically less susceptible to LD-induced confounding as compared with higher frequency variants; and though underpowered for the AMD case-control analyses, we expected them to have sufficient power for these quantitative trait analyses. We note that this additional MAF threshold was not applicable to *CFH*, which only included MAF<0.01% variants.

We first examined burden-ACAT p-values summarizing evidence for the association of each gene with each of the three OCT phenotypes. Setting a Bonferroni-corrected significance threshold of p < 0.05/12 = 0.0041 (3 OCT phenotypes * 2 genes * 2 adjustment models), we observe both *CFHR5* and *CFH* to significantly associate with outer segment of the photoreceptor layer across both adjustment models (more rigorous adjustment model: *CFHR5* p=0.0014, *CFH* p=0.0011), whereas neither gene displays nominal associations with inner segment of the photoreceptor layer or macular thickness at the inner inferior subfield (**Table S15**).

### CFHR5 ultra-rare variants contribute to AMD-protective changes in the photoreceptor layer

We examined the *CFHR5* and *CFH* individual variant set associations with outer segment of the photoreceptor layer, setting a Bonferroni-corrected significance threshold of p < 0.05/12 = 0.0041. When adjusting for Y402H and *ΔCFHR3-CFHR1*, we see that *CFHR5* rare (MAF<1%) pLOFs combined with AlphaMissense-predicted-damaging missense variants significantly associate with increased thickness of the outer segment of the photoreceptor layer (+0.12, p=8.1×10^−4^), and this association remains suggestive (p=0.0085) in the more rigorously adjusted model (**Table 4**). Furthermore, we observe that this same *CFHR5* variant mask limited to ultra-rare (MAF<0.1%) variants significantly associates with increased photoreceptor layer outer segment thickness across both adjustment models, with larger effect estimates than observed for the MAF<1% variant sets (more rigorous adjustment model: +0.34 SD, p=4.0×10^−4^; **Figure 3**). Thus, when excluding the higher frequency *CFHR5* rare variants driving the association with AMD, we still observe significant associations with increased thickness of this AMD-predictive OCT phenotype. Analyses of *CFHR5* pLOFs aggregated on their own yield similar results. These observed associations, including for ultra-rare *CFHR5* variants, support *CFHR5* genetic inhibition as having a causal, beneficial impact on the AMD-associated outer segment of the photoreceptor layer (recall from **Table 3** that thinning of this phenotype is a predictor of AMD).

**Table 4.**
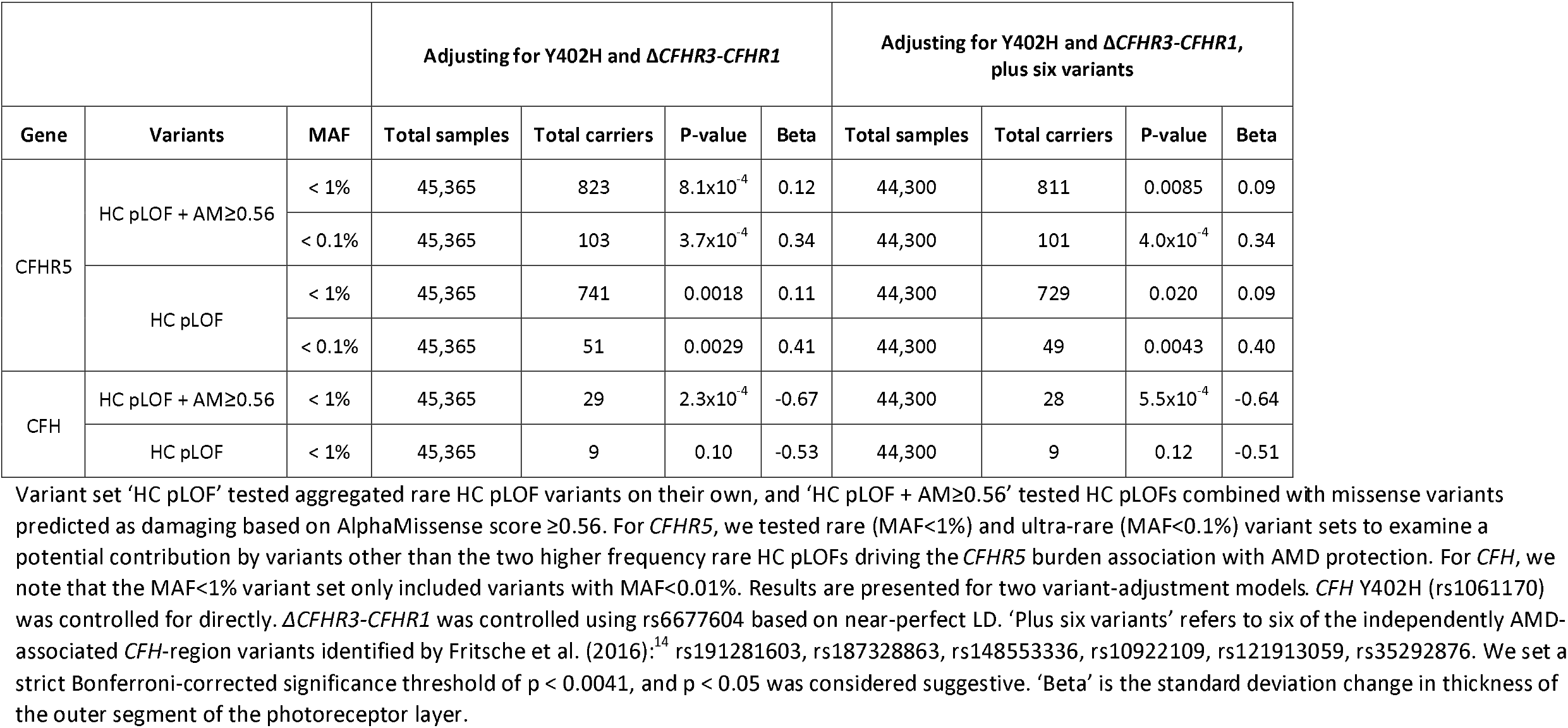
*CFHR5* and *CFH* rare (MAF<1%) predicted-damaging variant associations with thickness of the photoreceptor layer outer segment.

**Figure 3.**
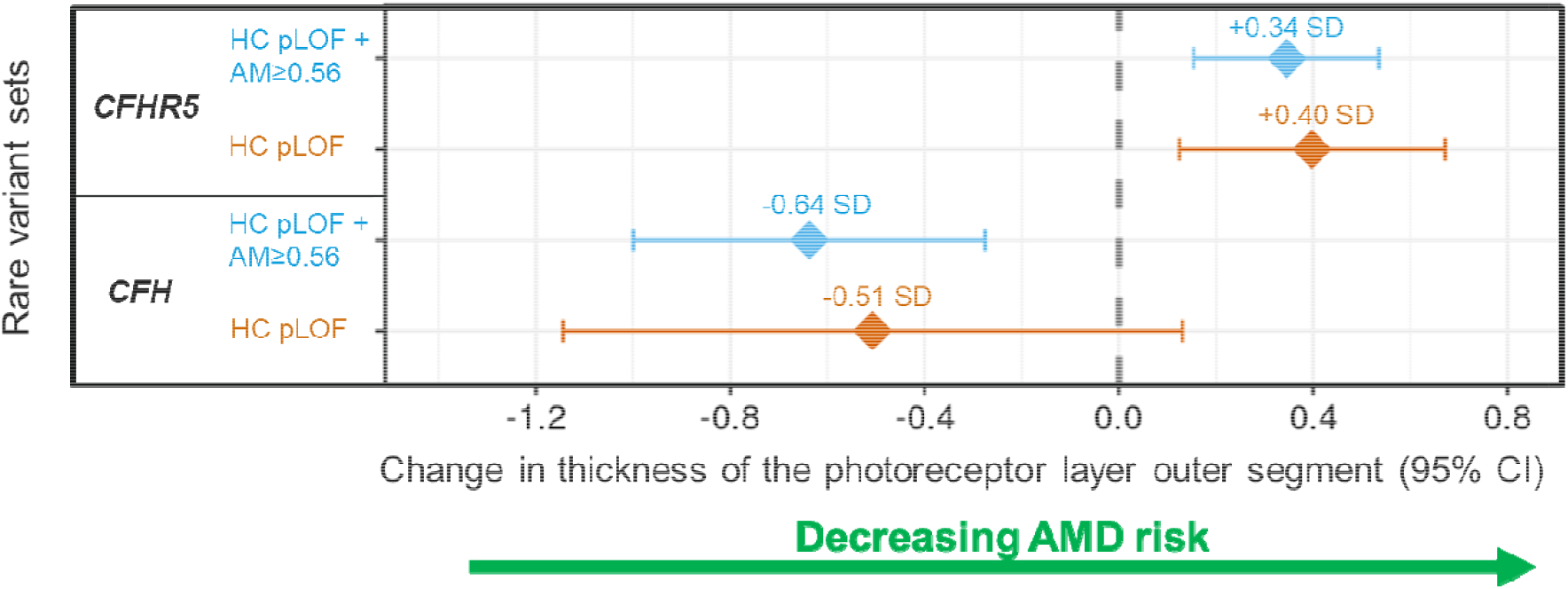
Rare predicted-damaging variants in *CFHR5* and *CFH* significantly associate with photoreceptor layer thickness (outer segment) in opposite directions. Shown are associations of *CFHR5* and *CFH* rare variant burden with thickness of the outer segment of the photoreceptor layer (measured at the central subfield), for the more rigorous model adjusting for *CFH* Y402H, *ΔCFHR3-CFHR1*, and several additional independently AMD-associated *CFH*-region variants. Results are plotted for HC pLOFs aggregated on their own (‘HC pLOF’) and in combination with AlphaMissense-predicted-damaging missense variants (‘HC pLOF + AM≥0.56’). *CFHR5* results are for the MAF<0.1% variant sets, and *CFH* results are for the MAF<1% sets (which only include ultra-rare variants). Increased thickness of the photoreceptor layer outer segment corresponds with decreased AMD risk (**Table 3; Table S13**). ‘SD’ is standard deviation. ‘CI’ is confidence interval.

For *CFH*, we see that rare pLOFs combined with AlphaMissense-predicted-damaging missense variants significantly associate with decreased thickness of the photoreceptor layer outer segment in both adjustment models (more rigorous adjustment model: -0.64 SD, p=5.5×10-4; **Table 4; Figure 3**). The test of *CFH* pLOFs on their own included only nine carriers and was underpowered to detect an association.

We observe similar association estimates when additionally controlling for the one remaining independently AMD-associated SNP at the *CFH* region identified by Fritsche et al. In these supplemental analyses, burden of rare predicted damaging variants in *CFHR5* and in *CFH* associates suggestively with increased (+0.31 SD, p=0.0049) and decreased (-0.68 SD, p=0.0044) thickness of the outer segment of the photoreceptor layer, respectively, despite a 27% reduction in analytic sample size due to high missingness of the additionally adjusted SNP (**Table S16**).

In combination with the observed OCT phenotype associations with incident AMD, these genetic associations observed when excluding all AMD cases lend further support to the notion that changes in the photoreceptor appear to precede AMD diagnosis. For *CFHR5* and *CFH*, supplemental analyses including the AMD cases yield nearly identical associations with increased and decreased thickness of the outer segment of the photoreceptor layer, respectively (**Table S17**). Supplemental investigation of *CFHR5* and *CFH* rare burden associations with all 40 OCT measures shows significant associations only for the photoreceptor layer outer segment thickness measures (**Table S18**).

As with AMD, we tested for a potential interaction between *CFH* Y402H and *CFHR5* rare predicted-damaging variants using photoreceptor layer outer segment thickness as the outcome, though we were not able to detect interaction (**Table S19**).

## DISCUSSION

In this work, we focused on clarifying potential roles for each *CFH*-region gene in AMD, with a particular focus on the *CFHR* genes. We used large-scale WES-based genetic and phenotypic data from the UKB study to test gene-based associations of rare (MAF<1%) predicted-damaging variants (including HC pLOFs and predicted-damaging missense variants) with AMD. We also used quantitative OCT measurements available for a subset of UKB participants, first identifying the specific OCT measures most strongly associated with AMD, then investigating rare variant associations with these top OCT predictors in tests that excluded all AMD cases. These analyses yielded novel findings linking rare pLOFs and predicted-damaging missense variants in *CFHR5* with improved outcomes for AMD and AMD-predictive retinal layer changes. Specifically, when controlling for *CFH*-region variants previously established to independently associate with AMD, we found that *CFHR5* rare variant burden significantly associates with decreased risk of AMD, with this association being primarily driven by pLOF variants, and appearing to be stronger for *CFH* Y402H risk allele carriers. In addition, we identified increased thinning of the outer segment of the photoreceptor layer of the retina to be a strong predictor of incident AMD and found *CFHR5* rare variant burden to significantly associate with increased thickness of this retinal layer, including when restricting to ultra-rare (MAF<0.1%) variants.

In testing rare variant burden associations with broadly defined AMD (ICD-10 H35.3), we identified significant associations for both *CFHR5* and *CFH*. For *CFHR5*, we observed significant AMD-protective associations for pLOFs aggregated on their own (OR=0.76, p=0.0017) and in combination with predicted-damaging missense variants (OR=0.75, p=7.1×10^−4^) when adjusting for Y402H and *ΔCFHR3-CFHR1*, and these associations remained suggestive after controlling for several additional AMD-associated variants at the *CFH* region (p<0.02). To our knowledge, this observation of a *CFHR5* rare pLOF-driven protective association with AMD that is demonstrated to be independent of nearby AMD signals has not previously been reported. It represents key evidence supporting a causal link between *CFHR5* genetic inhibition and decreased AMD risk and an important addition to prior research that has suggested a possible role for *CFHR5* in AMD.^14,15,47,48^ Follow-up examination revealed that one of the two *CFHR5* pLOFs driving the AMD-protective association (rs565457964-CAA) has a several-fold higher frequency among individuals with Finnish genetic ancestry and significantly associates with reduced risks for both dry and wet AMD in the Finnish population, supporting our rare variant findings.

We also observed evidence consistent with the *CFHR5* rare burden association with AMD protection being greater for *CFH* Y402H risk allele carriers (interaction p=0.04). Such interaction seems plausible, as the CFHRs have been shown to compete with CFH for binding to various surfaces (including C3b).^49,50^ Individuals with elevated AMD risk due to possible decreased binding activity of CFH to surfaces caused by *CFH* Y402H^51-53^ may derive greater benefit from *CFHR5* loss-of-function that yields a meaningful reduction in competition between CFHR5 protein and an already disadvantaged CFH.

For *CFH*, we observed burden of rare pLOFs on their own and combined with predicted-damaging missense variants to significantly associate with increased risk of broadly defined AMD across both adjustment models (pLOFs alone, more rigorous adjustment model: OR=6.95, p=2×10^−6^). This result aligns with expectations based on prior studies of *CFH* rare variants and AMD.^14,54,55^ We also found *CFH* rare predicted-damaging variants to significantly associate with increased risk for dry AMD, further supporting prior evidence linking the *CFH* region with dry AMD.^14^ Our analyses were underpowered to detect associations with wet AMD.

We identified thinner outer segment of the photoreceptor layer and thicker inner segment of the photoreceptor layer to be the strongest OCT phenotype predictors of both prevalent and incident AMD. The observation of photoreceptor layer thickness as the strongest predictor of AMD is consistent with findings from Zekavat et al. (2021).^16^ However, Zekavat et al. tested a photoreceptor layer thickness phenotype that combined the outer and inner segment photoreceptor layer measures, preventing identification of these measures as having independent AMD associations in opposite directions. Our findings are consistent with prior research showing that AMD polygenic risk score significantly associates with thinner photoreceptor layer outer segment and thicker photoreceptor layer inner segment.^17^ We then tested *CFHR5* and *CFH* rare variant burdens for association with these top AMD-predictive OCT phenotypes, excluding all AMD cases to enable analyses that were semi-orthogonal to our AMD case-control analyses. Across both adjustment models, we observed significant associations of *CFHR5* and *CFH* rare burden with increased and decreased thickness of the photoreceptor layer outer segment, respectively, consistent with our findings of these genes having opposite directions for association with AMD. For *CFHR5*, we see that ultra-rare (MAF<0.1%) predicted-damaging variants contribute significantly to increased thickness of the outer segment of the photoreceptor layer, further strengthening support for a causal role of *CFHR5* in AMD and AMD-associated phenotypes. We did not observe rare burden associations with inner segment of the photoreceptor layer, suggesting that the photoreceptor layer outer segment may be more relevant to the association of *CFH*-region genetic variants with AMD risk.

In contrast to the evidence for interaction between *CFHR5* rare variants and *CFH* Y402H in the AMD case-control analyses, we observed no evidence suggesting that the association of *CFHR5* rare variant burden with photoreceptor layer outer segment thickness may differ by Y402H genotype. Though this may appear at odds with the possibility of *CFHR5* rare variant protective associations with AMD being greater for Y402H risk allele carriers, a potential explanation may be that *CFHR5* rare variants associate with the same amount of increased thickness in photoreceptor layer outer segment regardless of Y402H carrier status, but this same increased thickness has the greatest beneficial impact for those at heightened AMD risk due to carrying the *CFH* Y402H allele.

Considering low carrier counts, our analyses were underpowered to effectively examine associations of AMD and AMD-related OCT traits with rare variant burdens in *CFHR1, CFHR2, CFHR3* and *CFHR4*. Thus, we cannot draw conclusions regarding a role or lack thereof for these genes in AMD. Prior studies have suggested a possible role for each of these other *CFHR* genes in AMD,^12,13,15,56^ and larger samples sizes will enable well-powered rare burden analyses to help clarify these potential roles.

With the exception of *CFH*, we had limited power to detect associations of aggregated rare variants with strictly defined AMD (dry and wet AMD). We were able to detect such associations for *CFH* because rare pLOFs and predicted-damaging missense variants in *CFH* yield very large risk-increasing associations with AMD. More moderately sized associations in a protective direction, as we would expect for *CFHR5* considering our results, have a low likelihood of being detected using only a few hundred strictly defined AMD cases. To overcome this power limitation, we analyzed a broadly defined AMD phenotype (ICD-10 H35.3), which evidence supports as being a relevant generalized phenotype for gaining insight into AMD. We also analyzed quantitative OCT measures that we found to be strong predictors of strictly defined AMD. Future studies will benefit from a larger collection of AMD-specific cases to enable better powered examination of dry and wet AMD-specific associations with rare pLOF and predicted-damaging missense variants, and to clarify and compare top OCT phenotype predictors for dry and wet AMD subtypes. We note that analyses of common variants at the *CFH* region, including the FinnGen analyses of *CFHR5* pLOF rs565457964-CAA, have identified similar associations with both dry and wet AMD.^14,44,45^

An important limitation of our study is that analyses were restricted to individuals with European genetic ancestry. Performing separate analyses for different ancestral groupings is a standard statistical genetics approach that helps reduce bias that can arise due to confounding by genetic ancestry. The UKB study overwhelmingly consists of participants with European ancestry, and we therefore focused our analyses on this group and excluded other ancestral groupings that were very underpowered for gene-based rare variant analyses. Future studies are needed that include large sample sizes of non-European genetic ancestry groupings with sequencing-based genotype data to allow well-powered rare variant analyses in these other populations. This will enable determination of the extent to which associations of certain genes with AMD may differ across populations, which is important for genetically informed drug discovery that benefits all individuals.

In conclusion, we have presented a collection of evidence that strongly supports genetic inhibition of *CFHR5* as protecting against AMD. These findings have the potential to inform the development of more effective therapeutics for AMD, including both dry and wet subtypes.

## Data Availability

Individual-level data analyzed in this study are not public due to privacy restrictions but are available to qualified investigators by application to the UK Biobank. All data generated by these analyses are contained in the published article and the supplemental data.

## APPENDIX A

### Validation of *CFHR5* and *CFH* rare variant sets using proteomics data

As about 10% of UKB participants have Olink-based proteomics data, we were able to use these data to examine the effect on protein levels for our gene-based rare variant sets. We used burden testing to analyze each *CFHR5* and *CFH* rare (MAF<1%) variant set for association with serum protein levels of CFH, CFHR2, CFHR4 and CFHR5 (measurements for CFHR1 and CFHR3 were not available). Results are provided in **Table S2**.

When adjusting for *CFH* Y402H and *ΔCFHR3-CFHR1*, the *CFHR5* variant set combining HC pLOFs with AlphaMissense-predicted-damaging missense variants (the set most strongly associated with decreased risk of broadly defined AMD) is very strongly associated with decreased CFHR5 protein (-1.70 SD, -log10p=507.2), with significant associations also seen with CFHR2 (-0.46 SD, p=1.1×10^−45^) and CFHR4 (-0.35 SD, p=5.3×10^−24^) but not with CFH (-0.007 SD, p=0.85) (**Table S2**). In the more rigorously adjusted model controlling for other AMD signals in the *CFH* region, the association with decreased CFHR5 protein remains very strong (-1.66 SD, -log10p=489.8) while associations with protein levels for CFHR2 (-0.15 SD, p=8×10^−7^) and CFHR4 (-0.04 SD, p=0.20) become strongly attenuated. Furthermore, in the most rigorous model that also controls for *CFH*-region SNP rs61818925 (resulting in a 27% reduction in analytic sample size), the very strong association with decreased CFHR5 protein is still observed (-1.62 SD, -log10p=392.9) while the association with CFHR2 protein becomes non-significant (-0.08 SD, p=0.008). The same results are observed for burden tests of *CFHR5* HC pLOFs on their own. In addition, when testing the *CFHR5* variant sets for association with all ∼3,000 Olink assayed proteins in models unadjusted for other variants, these rare variant sets are all by far most strongly associated with CFHR5 serum protein levels; and aside from the aforementioned associations with other *CFH*-region proteins, they also significantly but far less strongly associate with serum protein levels for F13B, CDHR5, CLNS1A and CFP (**Table S20**).

For the analyses of *CFHR5* rare burden associations with OCT phenotypes, we additionally tested variant sets limited to ultra-rare MAF<0.1% variants. When adjusting for Y402H and *ΔCFHR3-CFHR1*, the set of ultra-rare *CFHR5* HC pLOFs with AlphaMissense-predicted-damaging missense variants associates most strongly with reduced CFHR5 protein (-1.78 SD, p=9.8×10^−72^), and also significantly associates with CFHR2 (-0.47 SD, p=4.0×10^−7^) but not with CFHR4 (+0.15 SD, p=0.12) or CFH (-0.22 SD, p=0.03) (**Table S21**). Results for the more rigorously adjusted model are very similar. In the most rigorous model that also controls for *CFH*-region SNP rs61818925, the association of *CFHR5* ultra-rare predicted-damaging variants remains very strong (-1.73 SD, p=1.1×10^−52^) while the association with CFHR2 is no longer significant (-0.22 SD, p=0.02). Results are similar for *CFHR5* ultra-rare HC pLOFs aggregated on their own.

In *CFH* rare burden analyses adjusted for Y402H and *ΔCFHR3-CFHR1*, we see that HC pLOFs combined with AlphaMissense-predicted damaging missense variants (the set most strongly associated with increased risk of broadly defined AMD) yield the strongest association with CFH serum protein, significantly associating with decreased CFH protein (-1.55 SD, p=5.9×10^−21^). This variant set also displays nominal association with decreased CFHR4 serum protein (p=0.004) and no association with protein levels for CFHR5 or CFHR2 (p>0.05). The same pattern of protein associations is observed for the *CFH* variant set that includes HC pLOFs only. These associations are largely unchanged when implementing the more rigorous variant-adjustment models. We further note that, when testing the *CFH* variant sets for association with all ∼3,000 Olink assayed proteins in models unadjusted for other variants, these two CFH variant sets are both most strongly associated with CFH serum protein, with the CFH HC pLOFs combined with AlphaMissense-predicted-damaging missense variants also associating with decreased CFB protein at a Bonferroni-corrected significance threshold (-0.97 SD, p=2×10^−8^) (**Table S20**).

We were unable to examine rare variant set associations with serum protein levels for CFHR1 and CFHR3, as these proteins were not included on the Olink assay. However, were able to access summary statistics from deCODE genetics’ study of genetic associations with 4,907 serum proteins assayed using the SomaLogic platform.^57^ This study was performed in >35,000 Icelanders, and the assayed proteins include CFH and all five CFHRs. Individual variant associations are available, including for the *CFHR5* HC pLOFs rs565457964-CAA and rs565457964-CA, which we observed to be driving the association of *CFHR5* predicted damaging variants with AMD protection. For rs565457964-CAA, by far the strongest association across all assayed proteins is with decreased CFHR5 protein (beta=-1.46, p=7.7×10^−43^). The only other Bonferroni-significant (p < 0.05/4907) rs565457964-CAA associations are with decreased CFHR2 (beta=-0.65, p=1.4×10^−10^), increased NDUFS4 (beta=+0.59, p=2.7×10^−10^), and decreased CFHR4 (beta=-0.58, p=5.2×10^−8^); there were no associations of rs565457964-CAA with protein levels for CFH, CFHR1 and CFHR3 (all have p>0.05). For rs565457964-CA, the only Bonferroni-significant association was with decreased serum CFHR5 (beta=-2.03, p=3.8×10^−16^); nominal associations were observed with decreased CFHR2 (beta=-0.90, p=1.4×10^−4^) and decreased CFHR4 (beta=-0.62, p=0.01), and there were no associations with CFH, CFHR1 or CFHR3 protein levels (all have p>0.05). Though it is possible that the common deletion *ΔCFHR3-CFHR1* has some impact on ability to assay the CFHR1 and CFHR3 proteins, this concern may be decreased by the observation that *ΔCFHR3-CFHR1* (more specifically, the tag SNP rs6677604) does significantly associate with decreased levels for both of these proteins.

These rare burden and individual genetic variant associations with Olink- and SomaLogic-assayed proteins increase confidence that the *CFHR5* and *CFH* rare variant sets display good specificity in associating with CFHR5 and CFH proteins levels, respectively. For *CFHR5*, this is especially true when implementing the more rigorous variant adjustment model that conditions on CFH Y402H, *ΔCFHR3-CFHR1*, and several other AMD-associated variants in the *CFH* region. As described in the main text, for these more rigorous analyses we observe associations of *CFHR5* rare variant burden with protection from broadly defined AMD and beneficial changes in AMD-predictive OCT retinal phenotypes.

## APPENDIX B

### *CFHR5* rare variant burden association with AMD protection is not explained by LD with coding variants from nearby genes

Using the UKBB WES data, we identified all coding variants (as well as non-coding variants annotated to the 5’UTR or 3’UTR) in the window defined as 1 megabase (Mb) upstream of the *CFHR5* transcription start site to 1 Mb downstream of the *CFHR5* stop site. We restricted to variants with at least 25 alternate allele carriers. For the two HC pLOFs identified as the main drivers of the association of *CFHR5* rare predicted damaging variants with AMD protection (rs565457964-CAA and rs565457964-CA), we identified all variants in moderate to high LD at r^2^≥0.50 or D’≥0.80. rs565457964-CAA, which yielded the strongest individual variant association with AMD protection, exhibited moderate to high LD with 28 variants (none with r^2^≥0.50). rs565457964-CA exhibited moderate to high LD with 33 variants (only the CFHR5 missense variant rs41299613-C, discussed in the main text, had r^2^≥0.50 with rs565457964-CA). Together, the two *CFHR5* HC pLOFs show moderate to high LD with 36 unique coding or UTR variants. We excluded the *CFHR5* missense variant rs41299613-C, which is predicted as damaging based on AlphaMissense score = 0.93 and is included in our *CFHR5* rare burden set. *CFH* Y402H (rs1061170) was also among the variants due to having D’≥0.80 with both *CFHR5* HC pLOFs, and we excluded Y402H since we already accounted for it through inclusion as a covariate in our analyses. This left 34 variants, none with r^2^≥0.50.

We tested these 34 variants individually for association with broadly defined AMD, using the rigorously adjusted model that controls for Y402H, *ΔCFHR3-CFHR1*, and several additional *CFH*-region independently AMD-associated variants identified by Fritsche et al. (2016).^14^ Ten of the 34 variants yielded potentially meaningful associations with broadly defined AMD, which we defined leniently as having beta estimates of +/-0.05; these 10 variants included all of the variants yielding nominal (p<0.05) associations (**Table S22**). Of these 10 variants, five were annotated to have a most severe consequence of synonymous variant. Based on these functional annotations, we viewed these variants as less likely to explain the AMD signal as compared with the two *CFHR5* HC pLOF variants driving our observed association, and we therefore excluded these five variants from further consideration. This left five variants: two *CFH* missense variants, one *F13B* missense variant, one *ASPM* splice region variant, and a *CFHR5* 5’UTR variant (**Table S22**).

These five variants were included as additional covariates (along with *CFH* Y402H, *ΔCFHR3-CFHR1*, and additional independent AMD signals at the *CFH* region) in a burden test of the association of broadly defined AMD with rare (MAF<1%) *CFHR5* HC pLOF plus AlphaMissense-predicted-damaging missense variants. We find that this model that also includes several high LD variants yields OR=0.78 (p=0.0052), which is actually a slightly stronger result than obtained when not conditioning on the high LD variants (**Table S23**). Furthermore, in a model that also conditions on the Fritsche et al. (2016) AMD-associated *CFH*-region variant with 27% missingness in our analytic sample (which we excluded from our main analyses), we still observe a nominal association (OR=0.79, p=0.019) despite a 27% decrease in analytic sample size (**Table S23**). This persistent evidence of *CFHR5* rare predicted damaging variants associating with AMD protection after extensive correction for previously known *CFH*-region AMD signals and high-LD coding variants offers further evidence supporting *CFHR5* genetic inhibition as the causal mechanism giving rise to this protective association with AMD.

Separately, we performed additional sensitivity analyses testing *CFHR5* rare (MAF<1%) variant burden associations with broadly defined AMD, further adjusting for *CFH* rare variant burden of HC pLOFs combined with AlphaMissense-predicted-damaging missense variants (the *CFH* rare variant set most strongly associated with AMD). In the model adjusting for *CFH* Y402H, *ΔCFHR3-CFHR1*, and *CFH* rare variant burden, we see that *CFHR5* HC pLOFs on their own (OR=0.75, p=0.0013) and combined with AlphaMissense-predicted damaging missense variants (OR=0.75, p=5.6×10^−4^) remain associated with AMD protection, with no meaningful impact by the additional adjustment for *CFH* rare variant burden. The same is true for the more rigorously adjusted model that controls for *CFH* Y402H, *ΔCFHR3-CFHR1*, several additional *CFH*-region AMD-associated signals identified from Fritsche et al., and *CFH* rare variant burden, for which we see that *CFHR5* HC pLOFs on their own (OR=0.80, p=0.016) and combined with AlphaMissense-predicted damaging missense variants (OR=0.79, p=0.0079) remain associated with AMD protection.

## APPENDIX C

### Outer and inner segments of the photoreceptor layer independently associate with AMD

As the photoreceptor layer outer and inner segments are directly adjacent to one another, we wondered whether the observed significant findings for both layers may reflect correlation between the layers rather than independent associations with AMD. We find that measures for these outer and inner segments of the photoreceptor layer are moderately correlated. For instance, focusing on measures taken at the central subfield, we see correlations of r=-0.33 among all analyzed samples, r=-0.34 considering only broadly defined AMD cases, and r=-0.54 among strictly defined AMD cases (dry and wet combined). In supplemental tests examining association of each of these OCT phenotypes with AMD while controlling for the other OCT phenotype (e.g., testing photoreceptor layer outer segment thickness for association with AMD while conditioning on photoreceptor layer inner segment thickness), we see that associations attenuate but remain significant (**Table S24**), demonstrating that the outer and inner segments of the photoreceptor layer each have independent associations with AMD.

## ACKNOWLEDGEMENTS

This research has been conducted using the UK Biobank Resource (Applications 26041 and 65851). We would like to thank the participants and researchers of UK Biobank for creating an open-access resource. We thank the UK Biobank Exome Sequencing Consortium and UK Biobank for facilitating exome sequencing of participants and the UKB Pharma Proteomics Project for generating proteomics data. We acknowledge the participants and investigators of the FinnGen study, and thank the FinnGen team for making their results publicly available. Data management and analytics were performed using the REVEAL/SciDB translational analytics platform from Paradigm4.

## REFERENCES

1. Wong, W.L., Su, X., Li, X., Cheung, C.M., Klein, R., Cheng, C.Y., and Wong, T.Y. (2014). Global prevalence of age-related macular degeneration and disease burden projection for 2020 and 2040: a systematic review and meta-analysis. Lancet Glob Health 2, e106–116. 10.1016/S2214-109X(13)70145-1.

2. Fleckenstein, M., Schmitz-Valckenberg, S., and Chakravarthy, U. (2024). Age-Related Macular Degeneration: A Review. JAMA 331, 147–157. 10.1001/jama.2023.26074.

3. Rudnicka, A.R., Jarrar, Z., Wormald, R., Cook, D.G., Fletcher, A., and Owen, C.G. (2012). Age and gender variations in age-related macular degeneration prevalence in populations of European ancestry: a meta-analysis. Ophthalmology 119, 571–580. 10.1016/j.ophtha.2011.09.027.

4. Rudnicka, A.R., Kapetanakis, V.V., Jarrar, Z., Wathern, A.K., Wormald, R., Fletcher, A.E., Cook, D.G., and Owen, C.G. (2015). Incidence of Late-Stage Age-Related Macular Degeneration in American Whites: Systematic Review and Meta-analysis. Am J Ophthalmol 160, 85–93 e83. 10.1016/j.ajo.2015.04.003.

5. Wolf, A.T., Harris, A., Oddone, F., Siesky, B., Verticchio Vercellin, A., and Ciulla, T.A. (2022). Disease progression pathways of wet AMD: opportunities for new target discovery. Expert Opin Ther Targets 26, 5–12. 10.1080/14728222.2022.2030706.

6. Rofagha, S., Bhisitkul, R.B., Boyer, D.S., Sadda, S.R., Zhang, K., and Seven-Up Study Group (2013). Seven-year outcomes in ranibizumab-treated patients in ANCHOR, MARINA, and HORIZON: a multicenter cohort study (SEVEN-UP). Ophthalmology 120, 2292–2299. 10.1016/j.ophtha.2013.03.046.

7. Garg, A., Nanji, K., Tai, F., Phillips, M., Zeraatkar, D., Garg, S.J., Sadda, S.R., Kaiser, P.K., Guymer, R.H., Sivaprasad, S., et al. (2024). The effect of complement C3 or C5 inhibition on geographic atrophy secondary to age-related macular degeneration: A living systematic review and meta-analysis. Surv Ophthalmol 69, 349–361. 10.1016/j.survophthal.2023.11.008.

8. Warwick, A., and Lotery, A. (2018). Genetics and genetic testing for age-related macular degeneration. Eye (Lond) 32, 849–857. 10.1038/eye.2017.245.

9. Klein, R.J., Zeiss, C., Chew, E.Y., Tsai, J.Y., Sackler, R.S., Haynes, C., Henning, A.K., SanGiovanni, J.P., Mane, S.M., Mayne, S.T., et al. (2005). Complement factor H polymorphism in age-related macular degeneration. Science 308, 385–389. 10.1126/science.1109557.

10. Haines, J.L., Hauser, M.A., Schmidt, S., Scott, W.K., Olson, L.M., Gallins, P., Spencer, K.L., Kwan, S.Y., Noureddine, M., Gilbert, J.R., et al. (2005). Complement factor H variant increases the risk of age-related macular degeneration. Science 308, 419–421. 10.1126/science.1110359.

11. Edwards, A.O., Ritter, R., 3rd, Abel, K.J., Manning, A., Panhuysen, C., and Farrer, L.A. (2005). Complement factor H polymorphism and age-related macular degeneration. Science 308, 421–424. 10.1126/science.1110189.

12. Fritsche, L.G., Lauer, N., Hartmann, A., Stippa, S., Keilhauer, C.N., Oppermann, M., Pandey, M.K., Kohl, J., Zipfel, P.F., Weber, B.H., and Skerka, C. (2010). An imbalance of human complement regulatory proteins CFHR1, CFHR3 and factor H influences risk for age-related macular degeneration (AMD). Hum Mol Genet 19, 4694–4704. 10.1093/hmg/ddq399.

13. Hughes, A.E., Orr, N., Esfandiary, H., Diaz-Torres, M., Goodship, T., and Chakravarthy, U. (2006). A common CFH haplotype, with deletion of CFHR1 and CFHR3, is associated with lower risk of age-related macular degeneration. Nat Genet 38, 1173–1177. 10.1038/ng1890.

14. Fritsche, L.G., Igl, W., Bailey, J.N., Grassmann, F., Sengupta, S., Bragg-Gresham, J.L., Burdon, K.P., Hebbring, S.J., Wen, C., Gorski, M., et al. (2016). A large genome-wide association study of age-related macular degeneration highlights contributions of rare and common variants. Nat Genet 48, 134–143. 10.1038/ng.3448.

15. Lores-Motta, L., van Beek, A.E., Willems, E., Zandstra, J., van Mierlo, G., Einhaus, A., Mary, J.L., Stucki, C., Bakker, B., Hoyng, C.B., et al. (2021). Common haplotypes at the CFH locus and low-frequency variants in CFHR2 and CFHR5 associate with systemic FHR concentrations and age-related macular degeneration. Am J Hum Genet 108, 1367–1384. 10.1016/j.ajhg.2021.06.002.

16. Zekavat, S.M., Sekimitsu, S., Ye, Y., Raghu, V., Zhao, H., Elze, T., Segre, A.V., Wiggs, J.L., Natarajan, P., Del Priore, L., et al. (2022). Photoreceptor Layer Thinning Is an Early Biomarker for Age-Related Macular Degeneration: Epidemiologic and Genetic Evidence from UK Biobank OCT Data. Ophthalmology 129, 694–707. 10.1016/j.ophtha.2022.02.001.

17. Kaye, R.A., Patasova, K., Patel, P.J., Hysi, P., Lotery, A.J., and U. K. Biobank Eye and Vision Consortium (2021). Macular thickness varies with age-related macular degeneration genetic risk variants in the UK Biobank cohort. Sci Rep 11, 23255. 10.1038/s41598-021-02631-2.

18. Bycroft, C., Freeman, C., Petkova, D., Band, G., Elliott, L.T., Sharp, K., Motyer, A., Vukcevic, D., Delaneau, O., O’Connell, J., et al. (2018). The UK Biobank resource with deep phenotyping and genomic data. Nature 562, 203–209. 10.1038/s41586-018-0579-z.

19. Ko, F., Foster, P.J., Strouthidis, N.G., Shweikh, Y., Yang, Q., Reisman, C.A., Muthy, Z.A., Chakravarthy, U., Lotery, A.J., Keane, P.A., et al. (2017). Associations with Retinal Pigment Epithelium Thickness Measures in a Large Cohort: Results from the UK Biobank. Ophthalmology 124, 105–117. 10.1016/j.ophtha.2016.07.033.

20. Patel, P.J., Foster, P.J., Grossi, C.M., Keane, P.A., Ko, F., Lotery, A., Peto, T., Reisman, C.A., Strouthidis, N.G., Yang, Q., et al. (2016). Spectral-Domain Optical Coherence Tomography Imaging in 67 321 Adults: Associations with Macular Thickness in the UK Biobank Study. Ophthalmology 123, 829–840. 10.1016/j.ophtha.2015.11.009.

21. Sun, B.B., Chiou, J., Traylor, M., Benner, C., Hsu, Y.H., Richardson, T.G., Surendran, P., Mahajan, A., Robins, C., Vasquez-Grinnell, S.G., et al. (2023). Plasma proteomic associations with genetics and health in the UK Biobank. Nature 622, 329–338. 10.1038/s41586-023-06592-6.

22. Van Hout, C.V., Tachmazidou, I., Backman, J.D., Hoffman, J.D., Liu, D., Pandey, A.K., Gonzaga-Jauregui, C., Khalid, S., Ye, B., Banerjee, N., et al. (2020). Exome sequencing and characterization of 49,960 individuals in the UK Biobank. Nature 586, 749–756. 10.1038/s41586-020-2853-0.

23. Backman, J.D., Li, A.H., Marcketta, A., Sun, D., Mbatchou, J., Kessler, M.D., Benner, C., Liu, D., Locke, A.E., Balasubramanian, S., et al. (2021). Exome sequencing and analysis of 454,787 UK Biobank participants. Nature 599, 628–634. 10.1038/s41586-021-04103-z.

24. Szustakowski, J.D., Balasubramanian, S., Kvikstad, E., Khalid, S., Bronson, P.G., Sasson, A., Wong, E., Liu, D., Wade Davis, J., Haefliger, C., et al. (2021). Advancing human genetics research and drug discovery through exome sequencing of the UK Biobank. Nat Genet 53, 942–948. 10.1038/s41588-021-00885-0.

25. Danecek, P., Bonfield, J.K., Liddle, J., Marshall, J., Ohan, V., Pollard, M.O., Whitwham, A., Keane, T., McCarthy, S.A., Davies, R.M., and Li, H. (2021). Twelve years of SAMtools and BCFtools. Gigascience 10. 10.1093/gigascience/giab008.

26. Broad Institute. Picard Tools. http://broadinstitute.github.io/picard/.

27. Karczewski, K.J., Gupta, R., Kanai, M., Lu, W., Tsuo, K., Wang, Y., Walters, R.K., Turley, P., Callier, S., Baya, N., et al. (2024). Pan-UK Biobank GWAS improves discovery, analysis of genetic architecture, and resolution into ancestry-enriched effects. medRxiv, 2024.2003.2013.24303864. 10.1101/2024.03.13.24303864.

28. Chang, C.C., Chow, C.C., Tellier, L.C., Vattikuti, S., Purcell, S.M., and Lee, J.J. (2015). Second-generation PLINK: rising to the challenge of larger and richer datasets. Gigascience 4, 7. 10.1186/s13742-015-0047-8.

29. Purcell, S., Chang, C. PLINK. www.cog-genomics.org/plink/2.0/.

30. Anderson, C.A., Pettersson, F.H., Clarke, G.M., Cardon, L.R., Morris, A.P., and Zondervan, K.T. (2010). Data quality control in genetic case-control association studies. Nat Protoc 5, 1564–1573. 10.1038/nprot.2010.116.

31. Patterson, N., Price, A.L., and Reich, D. (2006). Population structure and eigenanalysis. PLoS Genet 2, e190. 10.1371/journal.pgen.0020190.

32. Tzoumas, N., Kavanagh, D., Cordell, H.J., Lotery, A.J., Patel, P.J., and Steel, D.H. (2022). Rare complement factor I variants associated with reduced macular thickness and age-related macular degeneration in the UK Biobank. Hum Mol Genet 31, 2678–2692. 10.1093/hmg/ddac060.

33. McLaren, W., Gil, L., Hunt, S.E., Riat, H.S., Ritchie, G.R., Thormann, A., Flicek, P., and Cunningham, F. (2016). The Ensembl Variant Effect Predictor. Genome Biol 17, 122. 10.1186/s13059-016-0974-4.

34. Karczewski, K.J., Francioli, L.C., Tiao, G., Cummings, B.B., Alfoldi, J., Wang, Q., Collins, R.L., Laricchia, K.M., Ganna, A., Birnbaum, D.P., et al. (2020). The mutational constraint spectrum quantified from variation in 141,456 humans. Nature 581, 434–443. 10.1038/s41586-020-2308-7.

35. Rentzsch, P., Witten, D., Cooper, G.M., Shendure, J., and Kircher, M. (2019). CADD: predicting the deleteriousness of variants throughout the human genome. Nucleic Acids Res 47, D886–D894. 10.1093/nar/gky1016.

36. Ioannidis, N.M., Rothstein, J.H., Pejaver, V., Middha, S., McDonnell, S.K., Baheti, S., Musolf, A., Li, Q., Holzinger, E., Karyadi, D., et al. (2016). REVEL: An Ensemble Method for Predicting the Pathogenicity of Rare Missense Variants. Am J Hum Genet 99, 877–885. 10.1016/j.ajhg.2016.08.016.

37. Cheng, J., Novati, G., Pan, J., Bycroft, C., Zemgulyte, A., Applebaum, T., Pritzel, A., Wong, L.H., Zielinski, M., Sargeant, T., et al. (2023). Accurate proteome-wide missense variant effect prediction with AlphaMissense. Science 381, eadg7492. 10.1126/science.adg7492.

38. Brandes, N., Goldman, G., Wang, C.H., Ye, C.J., and Ntranos, V. (2023). Genome-wide prediction of disease variant effects with a deep protein language model. Nat Genet 55, 1512–1522. 10.1038/s41588-023-01465-0.

39. Pejaver, V., Byrne, A.B., Feng, B.J., Pagel, K.A., Mooney, S.D., Karchin, R., O’Donnell-Luria, A., Harrison, S.M., Tavtigian, S.V., Greenblatt, M.S., et al. (2022). Calibration of computational tools for missense variant pathogenicity classification and ClinGen recommendations for PP3/BP4 criteria. Am J Hum Genet 109, 2163–2177. 10.1016/j.ajhg.2022.10.013.

40. Mbatchou, J., Barnard, L., Backman, J., Marcketta, A., Kosmicki, J.A., Ziyatdinov, A., Benner, C., O’Dushlaine, C., Barber, M., Boutkov, B., et al. (2021). Computationally efficient whole-genome regression for quantitative and binary traits. Nat Genet 53, 1097–1103. 10.1038/s41588-021-00870-7.

41. Liu, Y., Chen, S., Li, Z., Morrison, A.C., Boerwinkle, E., and Lin, X. (2019). ACAT: A Fast and Powerful p Value Combination Method for Rare-Variant Analysis in Sequencing Studies. Am J Hum Genet 104, 410–421. 10.1016/j.ajhg.2019.01.002.

42. R Core Team. (2021). R: A language and environment for statistical computing. (R Foundation for Statistical Computing).

43. Chen, S., Francioli, L.C., Goodrich, J.K., Collins, R.L., Kanai, M., Wang, Q., Alfoldi, J., Watts, N.A., Vittal, C., Gauthier, L.D., et al. (2024). A genomic mutational constraint map using variation in 76,156 human genomes. Nature 625, 92–100. 10.1038/s41586-023-06045-0.

44. Kurki, M.I., Karjalainen, J., Palta, P., Sipila, T.P., Kristiansson, K., Donner, K.M., Reeve, M.P., Laivuori, H., Aavikko, M., Kaunisto, M.A., et al. (2023). FinnGen provides genetic insights from a well-phenotyped isolated population. Nature 613, 508–518. 10.1038/s41586-022-05473-8.

45. FinnGen. (2024). Release DF11. r11.finngen.fi.

46. Wang, G., Sarkar, A., Carbonetto, P., and Stephens, M. (2020). A simple new approach to variable selection in regression, with application to genetic fine mapping. J R Stat Soc Series B Stat Methodol 82, 1273–1300. 10.1111/rssb.12388.

47. Sun, B.B., Kurki, M.I., Foley, C.N., Mechakra, A., Chen, C.Y., Marshall, E., Wilk, J.B., Biogen Biobank, T., Chahine, M., Chevalier, P., et al. (2022). Genetic associations of protein-coding variants in human disease. Nature 603, 95–102. 10.1038/s41586-022-04394-w.

48. Emilsson, V., Gudmundsson, E.F., Jonmundsson, T., Jonsson, B.G., Twarog, M., Gudmundsdottir, V., Li, Z., Finkel, N., Poor, S., Liu, X., et al. (2022). A proteogenomic signature of age-related macular degeneration in blood. Nat Commun 13, 3401. 10.1038/s41467-022-31085-x.

49. Alic, L., Papac-Milicevic, N., Czamara, D., Rudnick, R.B., Ozsvar-Kozma, M., Hartmann, A., Gurbisz, M., Hoermann, G., Haslinger-Hutter, S., Zipfel, P.F., et al. (2020). A genome-wide association study identifies key modulators of complement factor H binding to malondialdehyde-epitopes. Proc Natl Acad Sci U S A 117, 9942–9951. 10.1073/pnas.1913970117.

50. Goicoechea de Jorge, E., Caesar, J.J., Malik, T.H., Patel, M., Colledge, M., Johnson, S., Hakobyan, S., Morgan, B.P., Harris, C.L., Pickering, M.C., and Lea, S.M. (2013). Dimerization of complement factor H-related proteins modulates complement activation in vivo. Proc Natl Acad Sci U S A 110, 4685–4690. 10.1073/pnas.1219260110.

51. Clark, S.J., Perveen, R., Hakobyan, S., Morgan, B.P., Sim, R.B., Bishop, P.N., and Day, A.J. (2010). Impaired binding of the age-related macular degeneration-associated complement factor H 402H allotype to Bruch’s membrane in human retina. J Biol Chem 285, 30192–30202. 10.1074/jbc.M110.103986.

52. Ormsby, R.J., Ranganathan, S., Tong, J.C., Griggs, K.M., Dimasi, D.P., Hewitt, A.W., Burdon, K.P., Craig, J.E., Hoh, J., and Gordon, D.L. (2008). Functional and structural implications of the complement factor H Y402H polymorphism associated with age-related macular degeneration. Invest Ophthalmol Vis Sci 49, 1763–1770. 10.1167/iovs.07-1297.

53. Skerka, C., Lauer, N., Weinberger, A.A., Keilhauer, C.N., Suhnel, J., Smith, R., Schlotzer-Schrehardt, U., Fritsche, L., Heinen, S., Hartmann, A., et al. (2007). Defective complement control of factor H (Y402H) and FHL-1 in age-related macular degeneration. Mol Immunol 44, 3398–3406. 10.1016/j.molimm.2007.02.012.

54. Taylor, R.L., Poulter, J.A., Downes, S.M., McKibbin, M., Khan, K.N., Inglehearn, C.F., Webster, A.R., Hardcastle, A.J., Michaelides, M., Bishop, P.N., et al. (2019). Loss-of-Function Mutations in the CFH Gene Affecting Alternatively Encoded Factor H-like 1 Protein Cause Dominant Early-Onset Macular Drusen. Ophthalmology 126, 1410–1421. 10.1016/j.ophtha.2019.03.013.

55. Triebwasser, M.P., Roberson, E.D., Yu, Y., Schramm, E.C., Wagner, E.K., Raychaudhuri, S., Seddon, J.M., and Atkinson, J.P. (2015). Rare Variants in the Functional Domains of Complement Factor H Are Associated With Age-Related Macular Degeneration. Invest Ophthalmol Vis Sci 56, 6873–6878. 10.1167/iovs.15-17432.

56. Cantsilieris, S., Nelson, B.J., Huddleston, J., Baker, C., Harshman, L., Penewit, K., Munson, K.M., Sorensen, M., Welch, A.E., Dang, V., et al. (2018). Recurrent structural variation, clustered sites of selection, and disease risk for the complement factor H (CFH) gene family. Proc Natl Acad Sci U S A 115, E4433–E4442. 10.1073/pnas.1717600115.

57. Ferkingstad, E., Sulem, P., Atlason, B.A., Sveinbjornsson, G., Magnusson, M.I., Styrmisdottir, E.L., Gunnarsdottir, K., Helgason, A., Oddsson, A., Halldorsson, B.V., et al. (2021). Large-scale integration of the plasma proteome with genetics and disease. Nat Genet 53, 1712–1721. 10.1038/s41588-021-00978-w.

