## Supplemental Information for "Rare predicted loss-of-function and damaging missense variants in *CFHR5* associate with protection from age-related macular degeneration"

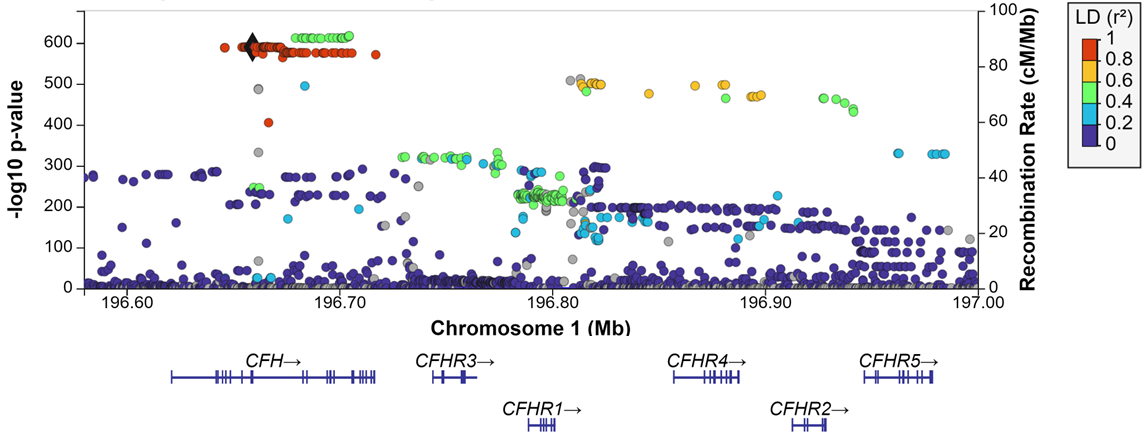

**Figure S1. LocusZoom plot of results from the Fritsche et al. (2016)^1^ AMD GWAS, for the CFH locus.** Generated at my.locuszoom.org. LD reference population is Eur. LD reference variant (black diamond) is *CFH* Y402H. URL to generate plot: <https://my.locuszoom.org/gwas/894486/region/?chrom=1&start=196580000&end=197000000&ld_variant=1%3A196659237_C%2FT>

**REFERENCES**

1.	Fritsche, L.G., Igl, W., Bailey, J.N., Grassmann, F., Sengupta, S., Bragg-Gresham, J.L., Burdon, K.P., Hebbring, S.J., Wen, C., Gorski, M., et al. (2016). A large genome-wide association study of age-related macular degeneration highlights contributions of rare and common variants. Nat Genet *48*, 134-143. 10.1038/ng.3448.
